# Is increased mortality by multiple exposures to COVID-19 an overseen factor when aiming for herd immunity?

**DOI:** 10.1101/2020.10.22.20217638

**Authors:** Kristina B. Helle, Arlinda Sadiku, Girma M. Zelleke, Aliou Bouba, Toheeb B. Ibrahim, H. Christian Tsoungui Obama, Vincent Appiah, Gideon A. Ngwa, Miranda I. Teboh-Ewungkem, Kristan A. Schneider

## Abstract

**Background:** Governments across the globe responded with different strategies to the COVID-19 pandemic. While some countries adapted draconic measures, which have been perceived controversial others pursued a strategy aiming for herd immunity. The latter is even more controversial and has been called unethical by the WHO Director-General. Inevitably, without proper control measure, viral diversity increases and multiple infectious exposures become common, when the pandemic reaches its maximum. This harbors not only a potential threat overseen by simplified theoretical arguments in support of herd immunity, but also deserves attention when assessing response measures to increasing numbers of infection.

**Methods and findings:** We extend the simulation model underlying the pandemic preparedness web interface CovidSim 1.1 (http://covidsim.eu/) to study the hypothetical effect of increased morbidity and mortality due to ‘multi infections’, either acquired at by successive infective contacts during the course of one infection or by a single infective contact with a multi-infected individual.

The simulations are adjusted to reflect roughly the situation in the East Coast of the USA. We assume a phase of general contact reduction (‘lockdown’) at the beginning of the epidemic and additional case-isolation measures. We study the hypothetical effects of varying enhancements in morbidity and mortality, different likelihoods of multi-infected individuals to spread multi infections and different susceptibility to multi infectious in different disease phases. It is demonstrated that multi infections lead to a slight reduction in the number of infections, as these are more likely to get isolated due to their higher morbidity. However, the latter substantially increases the number of deaths. Furthermore, simulations indicate that a potential second lockdown can substantially decrease the epidemic peak, the number of multi-infections and deaths.

**Conclusions:** Enhanced morbidity and mortality due to multiple disease exposure is a potential threat in the COVID-19 pandemic that deserves more attention. Particularly it underlines another facet questioning disease management strategies aiming for herd immunity.

## Introduction

The Coronavirus Disease 2019 (COVID-19) pandemic caused by the emergence of SARS Coronavirus 2 (SARS-CoV-2) infecting the respiratory system drastically changed the world in 2020. While most cases are asymptomatic or mild, severe infections require hospitalization or even ICU (intensive care unit) treatment, with symptoms of diffuse pneumonia, requiring oxygen supply and mechanical ventilation, potentially causing irreversible health damages or death by multiple organ failure [1].

There is evidence that inadequately protected front-line healthcare workers participated in the rapid initial spread of the pandemic [2], due to inadequate use of personal protective equipment (PPE) or reusing of single-use equipment, which potentially leads to multiple infectious contacts [3]. The mortality among healthcare workers, which is argued to be increased [4], received particular attention [5].

Governments across the globe reacted with different measures to manage the pandemic, many of them being controversial [6] and draconic. While some countries, initially did not strongly interfere with the pandemic, e.g., the UK, [7], or called out for voluntary social distancing, e.g., Sweden, others implemented draconic measures to mitigate the spread of the disease, e.g., Austria, New Zealand. These measures included lockdowns, cancellations of mass gatherings, mandatory social distancing, and isolation interventions that have been successively lifted. Indeed, some countries, e.g. Israel are calling out for a second lockdown, to address the so-called second wave.

The rationale behind lockdowns is to delay the pandemic to gain the time necessary, to build up healthcare and testing capacities, to develop efficient COVID-19 treatments, and save vaccinations. The latter allows us to immunize the population and avoid a full pandemic peak that would inevitably render the SARS-CoV-2 endemic. This would require constant adaptations of the vaccines as for influenza due to mutation in the virus.

On the contrary, the rationale behind herd immunity is to immunize the population naturally by not intervening with the pandemic [8] and not challenging the economy by draconic interventions. The argument of herd immunity polarizes with advocates in science [8] and politics [9] and also strict opponents[10]. Indeed, the WHO Director-General called the idea of herd immunity unethical [11].

The argument suggesting herd immunity often results from inadequate interpretations of oversimplified simulation models with a lack of in-depth knowledge of healthcare capacities and practices. Thus, overemphasizing the merits of herd immunity while downplaying its dangers. Typically, the implicit assumption of permanent immunity is made, but the decline of antibody levels and cases of COVID-19 reinfections suggest that immunity can be only temporal [12]. Moreover, elevated mortality due to shortages in health care resources including hospitals and ICUs is not accounted for and neither is the effect of permanent health damage and their derived long-term medical costs – for example medical costs for illnesses of the respiratory system in the Federal Republic of Germany amount to almost 5% of the healthcare costs in 2015 [13, Chapter 4]. Furthermore, indirect costs burdening the social system, e.g., expenses for working disabilities, are nethermost properly addressed. Importantly an uncontrolled pandemic spread will render the virus endemic facilitating its evolution and spread of novel variants that potentially attack the lung tissue more specifically like MERS [14].

An uncontrolled pandemic spread also facilitates the occurrence of multiple infectious exposures, which might increase the viral load within infections. Indeed, it has been argued that viral load correlates with disease severity, particularly in the context of mandatory usage of facial masks that could prevent from severe infections [15]. Increased viral load might be caused by multiple infectious exposures during the course of a COVID-19 episode.

Notably, multiple genetically distinguishable SARS-CoV-2 variants have been detected within different tissues of the same patient [16]. While the global genetic diversity of SARS-CoV-2 is large, the interactions between different viral variants within one organism are insufficiently understood [17]. Nevertheless, there is evidence that different viral variants differ in their contagiousness and virulence [17]. It should be mentioned that antibody-dependent enhancement was observed in corona viruses, which leads to more severe episodes due to repeated infectious contacts [18].

One of the current challenges in projecting mortality caused by COVID-19 is that estimates are based on data on the initial spread of the pandemic. During this phase, multiple infectious contacts during a disease episode are rare. If mortality is elevated in episodes caused by multiple infectious events with SARS-CoV-2 (super-infections), predictive models will underestimate this effect if not properly addressed. Remarkably, multiple infectious contacts during one episode are plausible for particular risk groups, such as inadequately protected healthcare workers [19], that are likely to be exposed to multiple infectious contacts within a short period of time and hence become super-infected.

Here, we introduce a mathematical model to study the effect of increased mortality due to the presence of multiple viral variants due to multiple infectious contacts (super-infections). The model extends the one from the freely available pandemic preparedness tool CovidSIM [20], which was also generalized in a very different way to study COVID-19 in closed facilities [21]. We study the effect of general contact reduction and case isolation measures on the extent of super-infections and its derived increased mortality. In particular, the effect of a second lockdown is investigated. The presentation follows the structure of [20] and [21].

## Methods

We study the hypothetical effect of multiple infections with COVID-19 during one infective period on disease severity and mortality. To do so we adapt the extended SEIR model underlying the pandemic preparedness tool CovidSIM [20]. Our deterministic compartmental model is illustrated in Fig 1. We give a verbal description of the model here and refer to S1 Appendix for a concise mathematical description.

**Fig 1.**
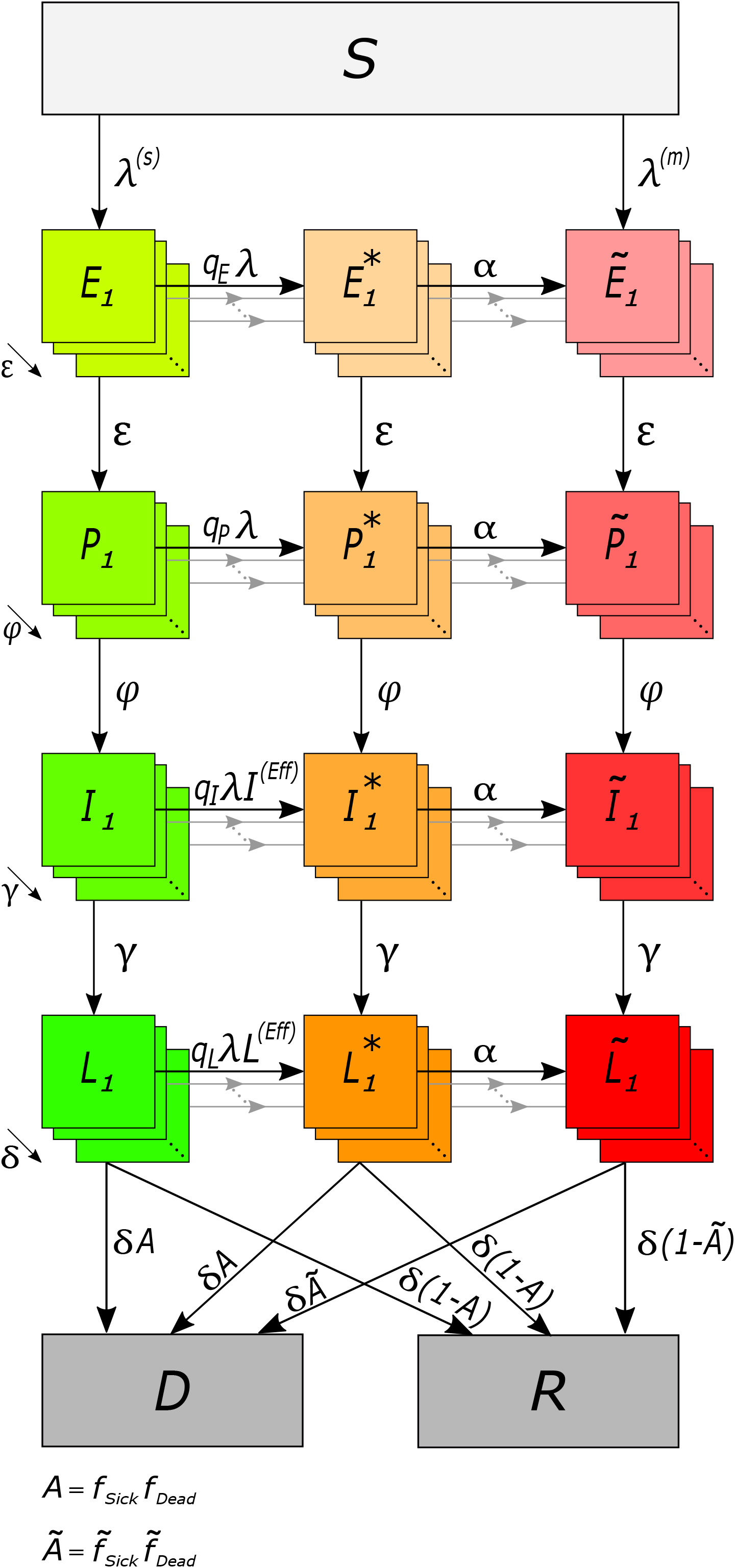
Model illustration. Model compartments are shown as boxes. Grey boxes illustrate susceptible (*S*), recovered (*R*), and dead (*D*) individuals. Green boxes illustrate single infections, orange boxes multi infections in the transient phase, and red boxes multi infections characterized by higher morbidity and mortality. Infected compartments consist of latent infected (*E*_*k*_), prodromal (*P*_*k*_), fully contagious (*I*_*k*_), and late infectious (*L*_*k*_) Erlang sub-states. Arrows indicate flows between compartments at the various rates.

A population of *N* individuals is assumed, subdivided into susceptible, infected, and recovered individuals. After a susceptible individual becomes infected it passes through the (i) latency period (the individual is not yet infectious), (ii) prodromal period (the individual does not yet exhibit characteristic symptoms, but is partly infectious), (iii) fully contagious period (the individual is either asymptomatic, or shows symptoms ranging from mild to severe), and (iv) the late infectious period (the individual is no longer fully contagious). After the late infectious phase individuals either recover (and become permanently immune) or they die. The model follows the time change of the numbers of individuals being susceptible (*S*), latently infected (*E*), prodromal (*P*), fully contagious (*I*), and late infectious (*L*), as well as those, who are recovered (*R*) or died (*D*). Deaths unrelated to COVID-19 are ignored, as we assume a pandemic in a large population in a relatively short time period.

A fraction of fully contagious individuals develops symptoms, i.e., they get sick. Asymptomatic infections always lead to recovery, while a fraction of symptomatic infections is lethal. We assume the fractions of symptomatic and lethal infections to be higher among individuals, which got infected multiple times with SARS-CoV-2 during the course of their infection. This reflects multiple viral variants acquired at a single infective event or at consecutive infective events and hence a higher viral load. We therefore follow individuals with ‘single’ and ‘multi’ infections throughout all phases of the infection. The numbers of individuals with multi infections in the various phases are denoted by 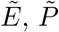, *Ĩ*, and 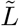. Individuals acquire a multi infection either at their initial infection, or through consecutive infective events during the course of their infection. The susceptibility to a second infection is different in the latent, prodromal, fully contagious, and late infectious phases. While at infective contacts single infected individuals spread only single infection, individuals with multi infections can spread single and multi infections.

Individuals that acquire a multi infection at their first infective contact can transmit multi infections, have a higher risk of developing symptoms, and have increased mortality as soon as they reach the respective phase of the disease. However, if individuals move from a single to a multi infection by a second infective contact, the characteristics of the multi infection manifest only after a time delay. During this time, individuals are moved into transient compartments *E*^*\**^, *P* ^*\**^, *I*^*\**^, and *L*^*\**^. As long as individuals are in the transient phases, they can neither spread multi infections nor are they more likely to show symptoms or die than individuals with single infections.

Susceptible individuals acquire infections through contacts with individuals in the prodromal, the fully contagious, or the late infectious periods at rates *β*_*P*_, *β*_*I*_, *β*_*L*_, respectively. Note, a fraction of multi-infected individuals transmits a multi infections, while the rest transmits only a single infection.

The basic reproduction number *R*_0_ is the average number of infections caused by a single infected individual in a susceptible population in which no intervention occurs during the whole infectious period, consisting of the prodromal, fully contagious, and late infectious phases. This quantity is assumed to fluctuate seasonally around its annual average 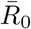.

The model incorporates general contact reduction measures as well as case isolation. General contact reduction, i.e., curfews or social distancing, reduces the contact rate between all individuals and will be sustained during a specific time interval. Regarding case isolation, a fraction of symptomatic cases (those that seek medical help) will be isolated in quarantine wards until recovery or death. (This fraction is higher in multi than in single infections, reflecting more severe symptoms and hence a higher demand for medical help.) If the wards are full, individuals are sent into home isolation. While perfect isolation is achieved in quarantine wards, home isolation is imperfect, i.e., not all infectious contacts are avoided. Also, case isolation mechanisms are sustained only during a specific time interval, and might not be in place at the introduction of the disease.

Classical SEIR models assume that individuals proceed from one infected stage to the next at rates being the reciprocal average residence time spent in each compartment. Implicitly, this assumes exponentially distributed residence times. This is undesirable [20, cf.], because (i) a proportion of individuals progresses too fast, whereas others progress too slow from one state to the next, and (ii) the variance of the average residence time in each compartment equals its mean. To model more realistic dynamics, we subdivide the latent, prodromal, fully contagious, and late infectious periods into several sub-stages, through which individuals pass successively. As a consequence, residence times follow Erlang-distributions, which are more realistic, because as the means of the residence times does not determine their variances.

### Implementation of the model

The model as described in S1 Appendix was numerically solved by a 4th order Runge-Kutta method. The code was implemented in Python 3.8 using the function solve ivp as part of the library Scipy, and library Numpy. Graphical output was created in Python 3.8. using library Matplotlib.

## Results

Model parameters are chosen to roughly reflect the situation in the United States of America (USA), where control measures were not as draconic as in other countries and a number of COVID-19 related deaths occurred among front line healthcare workers. The goal is to study the potential negative effects on morbidity and mortality due to multiple disease exposure during the epidemic peak – which is an overlooked risk factor in the debate about herd immunity.

S1 Table - S4 Table list the parameter values used in the numerical investigations. A population of *N* = 250 million (reflecting the US East Cost) was assumed. The first COVID-19 cases were confirmed in late February 2020 [19]. Because these initial cases correspond to imported cases from outside the population, time *t* = 0 corresponds to the middle of March 2020. To avoid confounding factors we simulated the dynamics with and without seasonal fluctuations in the basic reproduction number. Without seasonality *R*_0_ = 3.5 was assumed (this case serves as a baseline for comparisons). When incorporating seasonality, *R*_0_ fluctuated by *a* = 43% around its annual average 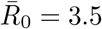 and peaked in late December 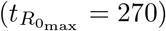. The average duration of the latent, prodromal, fully contagious and late infectious stages was assumed to be *D*_*E*_ = 3.7, *D*_*P*_ = 1, *D*_*I*_ = 7 and *D*_*L*_ = 7 days, respectively. In the prodromal and late infective stages, individuals were assumed to be half as infective as in the fully contagious stage. Multi-infected individuals were more likely to develop severe symptoms (default values: *f*_Sick_ = 58% *vs*. 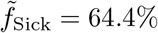) and had an increased mortality (default values: *f*_Dead_ = 3% *vs*. 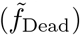 = 4%).

In the following, we describe the influence of the various model parameters characterizing multi infection

### Morbidity caused by multi infections

The increased fraction of developing symptoms in multi infections compared with single infections (higher 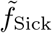 in comparison to *f*_Sick_) has three effects: (i) slightly fewer people get infected during the pandemic, because of case isolation. In particular, multi infections cause a higher proportion of symptomatic infections, which are isolated and no longer participate (fully) in transmission. The larger the fraction of multi infections that become symptomatic, the stronger is this effect. This manifests in higher number of susceptible infections (see Fig 2A, D). (ii) The height of the epidemic peak is slightly reduced as multi infections are more likely to be isolated and do not participate in disease transmission. Also, slightly fewer multi infections occur. This effect manifests during the onset of the epidemic peak, when multi infections become more frequent (see Fig 2B, E). (iii) The number of deaths clearly increases with the likelihood of having symptomatic infections, because a fraction of these are lethal (see Fig 2C, F).

**Fig 2.**
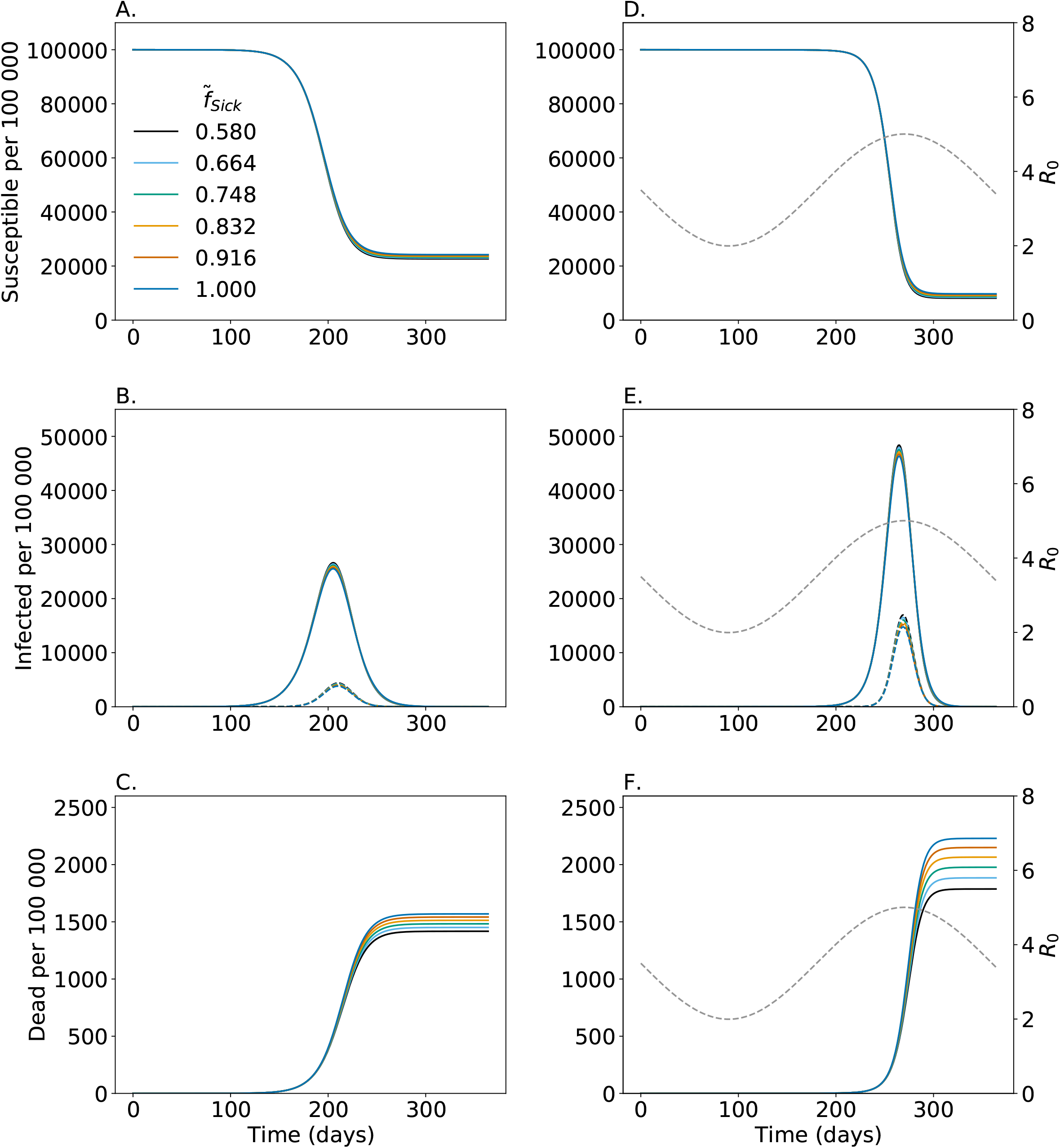
Proportion of multi infected individuals being symptomatic. Shown are the numbers of susceptibles (A, D); (B, E) infected consisting of all latent, prodromal, fully, and late contagious individuals among all single and multi infections (solid lines) and non-transient multi infections (dashed lines); and dead individuals (C, F). Colors are for different fractions of multi infected individuals that become symptomatic 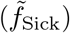. Figs A-C assume no seasonal fluctuations in *R*_0_, while D-F assume seasonal fluctuations in *R*_0_ illustrated by the gray dashed line corresponding to the y-axis on the right-hand side. Parameters are given in S1 Table – S4 Table.

Seasonality impacts the results (compare Figs 2A-C with 2D-F). If, as in the simulations, the epidemic peak is shifted into the onset of the flu season, more people get infected and the pandemic peak is higher and narrower. This leads to a disproportional higher number of multi infections due to seasonality in transmissibility. Particularly, the mortality is disproportionally increased (compare Figs 2C and F).

### Severity of symptoms

The severity of symptoms is reflected in the model by the fraction of individuals seeking medical help and become isolated. The more severe the symptoms of multi infections, the more of them get isolated (higher 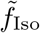). Isolating a higher fraction of symptomatic multi infections leads to slightly fewer infections and a reduced height of the epidemic peak (3A, D). Also, the peak number of multi infections decreases (3B, E). Moreover, there is a reduction in mortality (3C, F). However, this reduction does not compensate the potential increase in mortality due to super-infections (compare Figs 2C and F with 3C, F).

The same pattern applies in the absence and prescience of seasonality (compare Fig 3A-C with Fig 3D-F).

**Fig 3.**
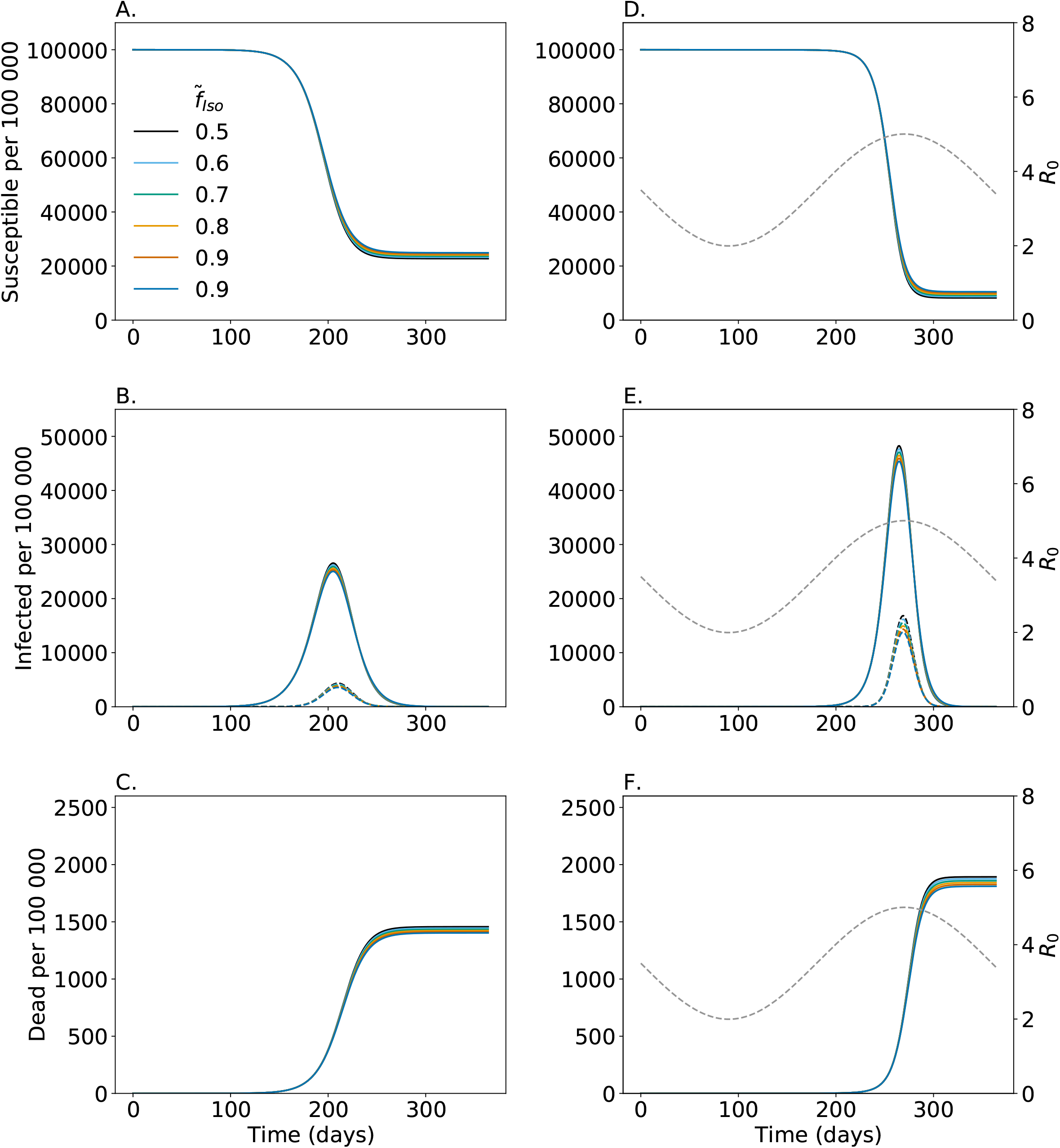
Proportion of symptomatic multi infections getting isolated. Shown are the numbers of susceptibles (A, D); (B, E) infected consisting of all latent, prodromal, fully, and late contagious individuals among all single and multi infections (solid lines) and non-transient multi infections (dashed lines); and dead individuals (C, F). Colors are for different fractions of symptomatic multi infected individuals with increased morbidity and mortality that become isolated 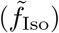. Figs A-C assume no seasonal fluctuations in *R*_0_, while D-F assume seasonal fluctuations in *R*_0_ illustrated by the gray dashed line corresponding to the y-axis on the right-hand side. Parameters are given in S1 Table – S4 Table.

### Increased mortality

As described above increased morbidity of multi infections reflected by lager fractions of symptomatic and isolated infections decreases the overall number of cases, but causes a higher number of deaths. This increase in mortality scales with the case fatality of multi infections reflected by the parameter 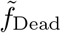. As illustrated by Fig 4 this parameter does not affect the number of infections, but the number of lethal infections. The higher case mortality of multi infections the higher the number of deaths. This effect is amplified by seasonal variation in *R*_0_ if the epidemic peak occurs in the flu season. Namely, the epidemic peak is higher, causing an excess in multi infections which are responsible for the increased mortality.

**Fig 4.**
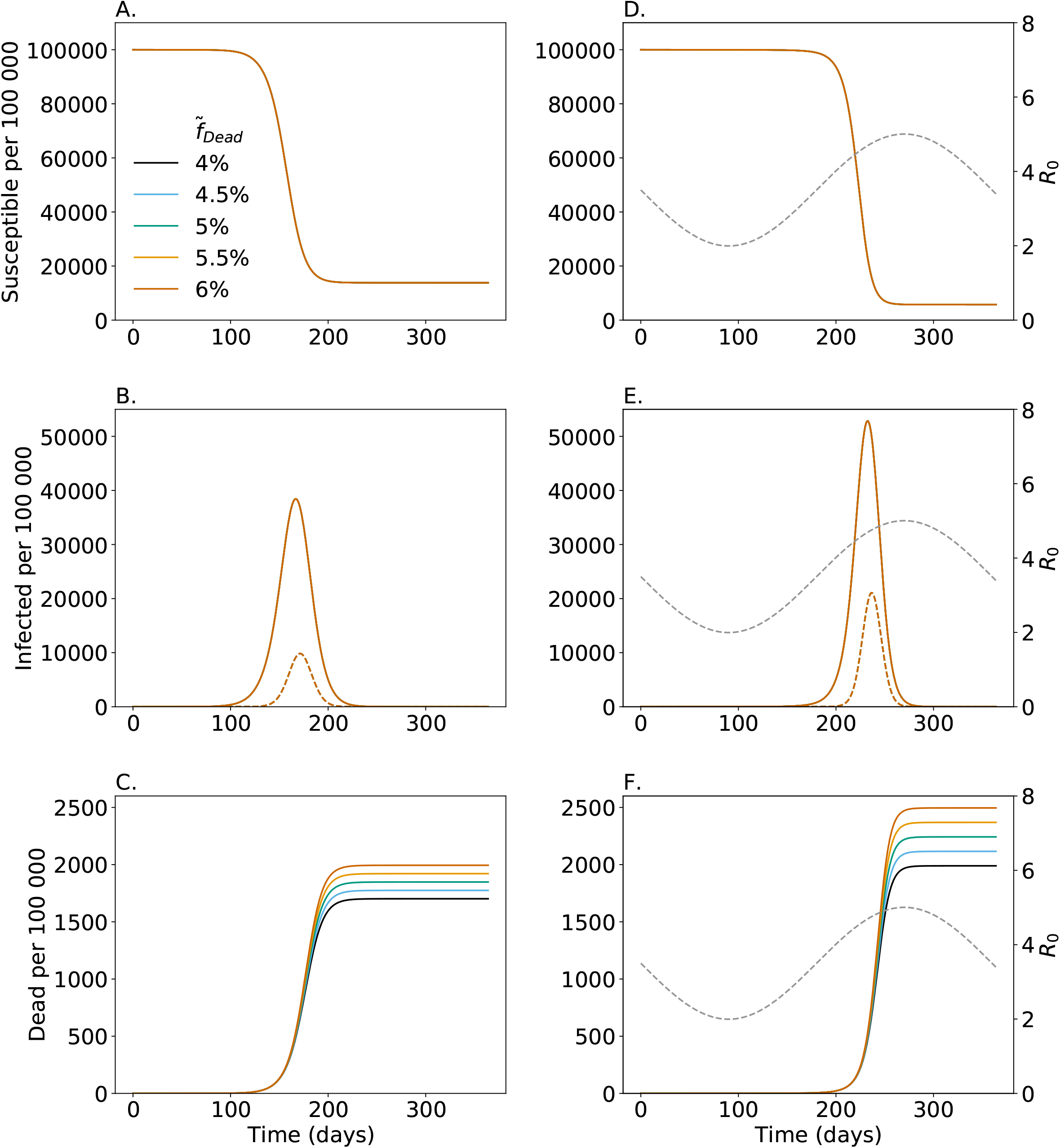
Increased mortality of symptomatic multi infections. Shown are the numbers of susceptibles (A, D); (B, E) infected consisting of all latent, prodromal, fully, and late contagious individuals among all single and multi infections (solid lines) and non-transient multi infections (dashed lines); and dead individuals (C, F). Colors are for different mortality of symptomatic multi infected individuals with increased morbidity and mortality 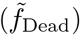. Figs A-C assume no seasonal fluctuations in *R*_0_, while D-F assume seasonal fluctuations in *R*_0_ illustrated by the gray dashed line corresponding to the y-axis on the right-hand side. Parameters are given in S1 Table – S4 Table.

### Likelihood of spreading multi infections

Multi infections are acquired either at a single infective event or at consecutive infective events. In case of the latter, the negative effects in terms of morbidity and mortality only manifest after a transient phase. In particular, in this phase individuals cannot spread multi infections and they might recover before the negative effects of multi infections are in place. The likelihood of multi infected individuals to spread multi infections 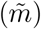 only marginally effects the number of infections during the epidemic (5A, B, D, E). However, naturally the peak number of multi infections is higher the larger 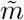. Thus, case fatality increases with 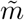.

The same results are obtained with and without seasonal fluctuations in *R*_0_ (compare Fig 5A-C with Fig 5D-F).

**Fig 5.**
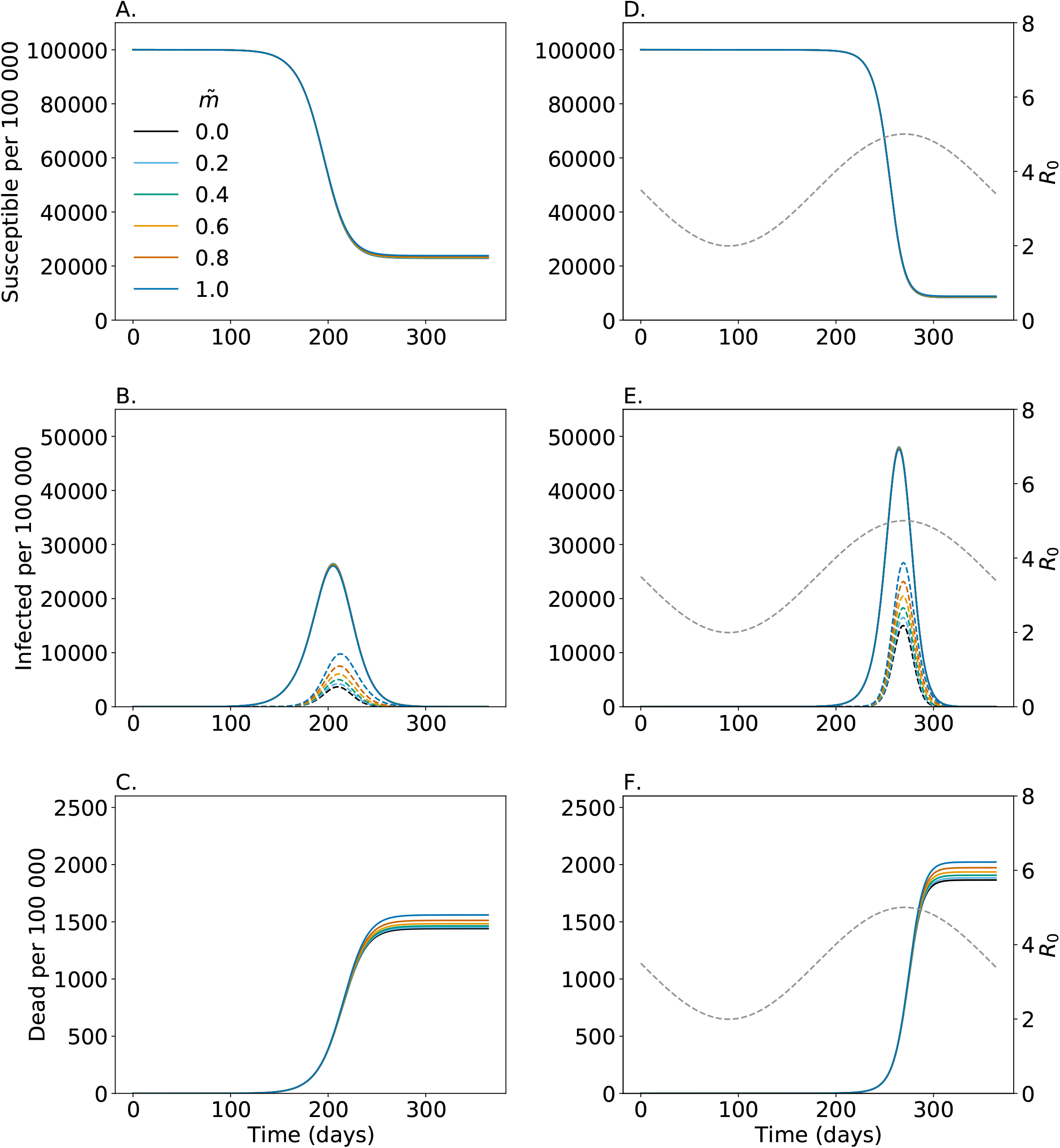
Likelihood to spread multi infections. Shown are the numbers of susceptibles (A, D); (B, E) infected consisting of all latent, prodromal, fully, and late contagious individuals among all single and multi infections (solid lines) and non-transient multi infections (dashed lines); and dead individuals (C, F). Colors are for different likelihoods of multi infected individuals to spread multi infections 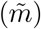. Figs A-C assume no seasonal fluctuations in *R*_0_, while D-F assume seasonal fluctuations in *R*_0_ illustrated by the gray dashed line corresponding to the y-axis on the right-hand side. Parameters are given in S1 Table – S4 Table.

### Susceptibility to successive infections

Individuals already infected with COVID-19 will show an immune reaction that mediates susceptibility to successive infections. Thus, the force of infection experienced by single infected individuals might be reduced compared with that acting on susceptible. Namely, only percentages *q*_*E*_, *q*_*P*_, *q*_*I*_, *q*_*L*_ of the force of infection are acting on single infected individuals in the latent, prodromal, fully contagious, and late infectious stages, respectively.

Typically, susceptibility to further infections should not be affected much in the latent phase, while it is strongly decreased in the late infectious phase, in which some immunity is already developed.

Clearly susceptibility to successive infections affects the number of multi infections. We studied the five scenarios listed in Table 1. The five scenarios correspond to decreasing susceptibility, with scenario (i) being the extreme that susceptibility is not affected, and scenario (iv) being the default used in the simulations here.

**Table 1.** Susceptibility to successive infections.

| Scenario | $q_E$ | $q_P$ | $q_I$ | $q_L$ |
| --- | --- | --- | --- | --- |
| i | 1 | 1 | 1 | 1 |
| ii | 1 | 1 | 1 | 0.75 |
| iii | 1 | 1 | 0.75 | 0.5 |
| iv | 1 | 0.75 | 0.5 | 0.25 |
| v | 0.75 | 0.5 | 0.25 | 0 |

The number of infections during the epidemic as well as the pandemic peak are almost indistinguishable for all scenarios (Fig 6A, D). However, the number of multi infections changes dramatically (Fig 6B, E). The reason is as follows: the number of multi infections is highest shortly after the epidemic peak when most infections are single infection, and it is thus more likely to get infected twice during the course of the disease than once by a multi infected individual. Reduced susceptibility of single infected individuals results in a clear reduction of multi infections. This reduced numbers of multi infections result in a reduced mortality (Fig 6C, F).

**Fig 6.**
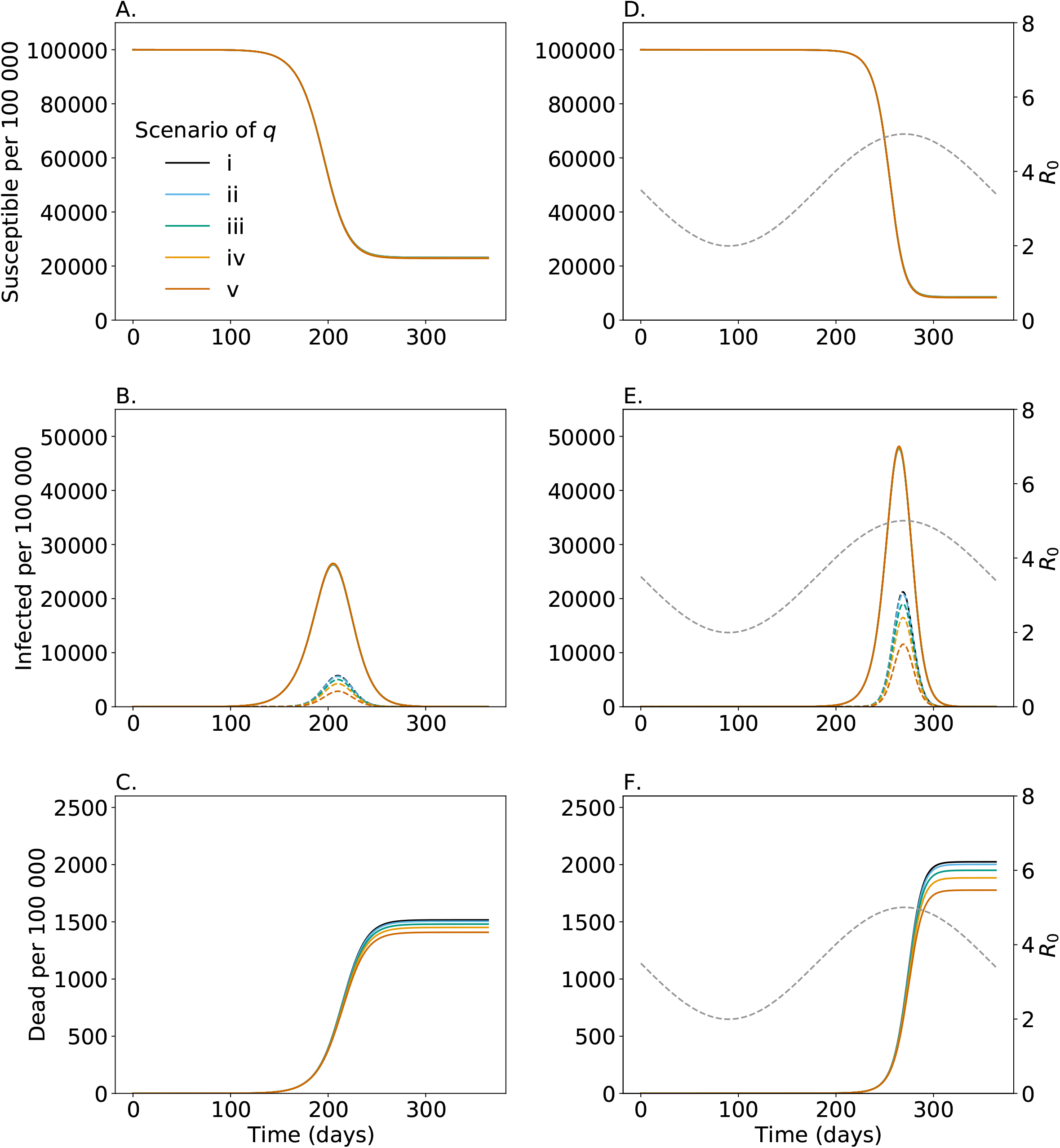
Susceptibility to successive infections. Shown are the numbers of susceptibles (A, D); (B, E) infected consisting of all latent, prodromal, fully, and late contagious individuals among all single and multi infections (solid lines) and non-transient multi infections (dashed lines); and dead individuals (C, F). Colors are the different scenarios of susceptibility during the various ineffective phases given in Table 1. Figs A-C assume no seasonal fluctuations in *R*_0_, while D-F assume seasonal fluctuations in *R*_0_ illustrated by the gray dashed line corresponding to the y-axis on the right-hand side. Parameters are given in S1 Table – S4 Table.

Comparisons of Figs 6A-C with Figs 6D-F changes show that seasonal fluctuations in *R*_0_ only quantitatively but not qualitatively affect the outcome.

### The effect of countermeasures and a second lockdown

The effects of general contact reduction and case isolation was studied for a simpler model in [20]. Here, we are interested in the effects of these measures on the number of multi infections and mortality. Moreover, we are interested in the effect of a second ‘lockdown’. More precisely, in a second phase of general contact reduction. We therefore study ten scenarios as listed in Table 2. Scenario 0 corresponds to no intervention and serves as a baseline comparison. All other scenarios assume case isolation but differ in general contact reduction. Scenarios (I)-(IV) assume an initial lockdown with different durations and effective contact reduction. They serve as comparison for evaluating the measures that were actually implemented. Scenarios (V)-(IX) illustrate the effect of a second (gentle) lockdown (with a reduction of 40% of contacts).

**Table 2.** Countermeasures.

| Scenario | $f_{\text{Iso}}$ | $t_{\text{Iso1}}$ | $t_{\text{Iso2}}$ | $p_{\text{Dist1}}$ | $t_{\text{Dist11}}$ | $t_{\text{Dist21}}$ | $p_{\text{Dist2}}$ | $t_{\text{Dist12}}$ | $t_{\text{Dist22}}$ |
| --- | --- | --- | --- | --- | --- | --- | --- | --- | --- |
| 0 | 0 | - | - | 0 | - | - | 0 | - | - |
| I | 0.4 | 0 | 365 | 0.4 | 0 | 45 | 0 | - | - |
| II | 0.4 | 0 | 365 | 0.4 | 0 | 85 | 0 | - | - |
| III | 0.4 | 0 | 365 | 0.6 | 0 | 45 | 0 | - | - |
| IV | 0.4 | 0 | 365 | 0.6 | 0 | 85 | 0 | - | - |
| V | 0.4 | 0 | 365 | 0.4 | 0 | 45 | 0.4 | 230 | 275 |
| VI | 0.4 | 0 | 365 | 0.4 | 0 | 45 | 0.4 | 240 | 285 |
| VII | 0.4 | 0 | 365 | 0.4 | 0 | 45 | 0.4 | 250 | 295 |
| VIII | 0.4 | 0 | 365 | 0.4 | 0 | 45 | 0.4 | 260 | 305 |
| IX | 0.4 | 0 | 365 | 0.4 | 0 | 45 | 0.4 | 270 | 355 |

The effects of the measures can be best understood in the absence of seasonal fluctuations in *R*_0_. However, this results in unrealistic dynamics (because the second lockdown might start too late). Hence, the results without seasonality serve only as a comparison.

Without seasonality, general contact reduction only delays the epidemic peak (Fig 7A, B). It does not reduce its height (Fig 7B), which is reduced by case isolation (compare scenario 0 with scenarios I-IV). Also, the number of multi infections and deaths do not change (Fig 7B, C).

**Fig 7.**
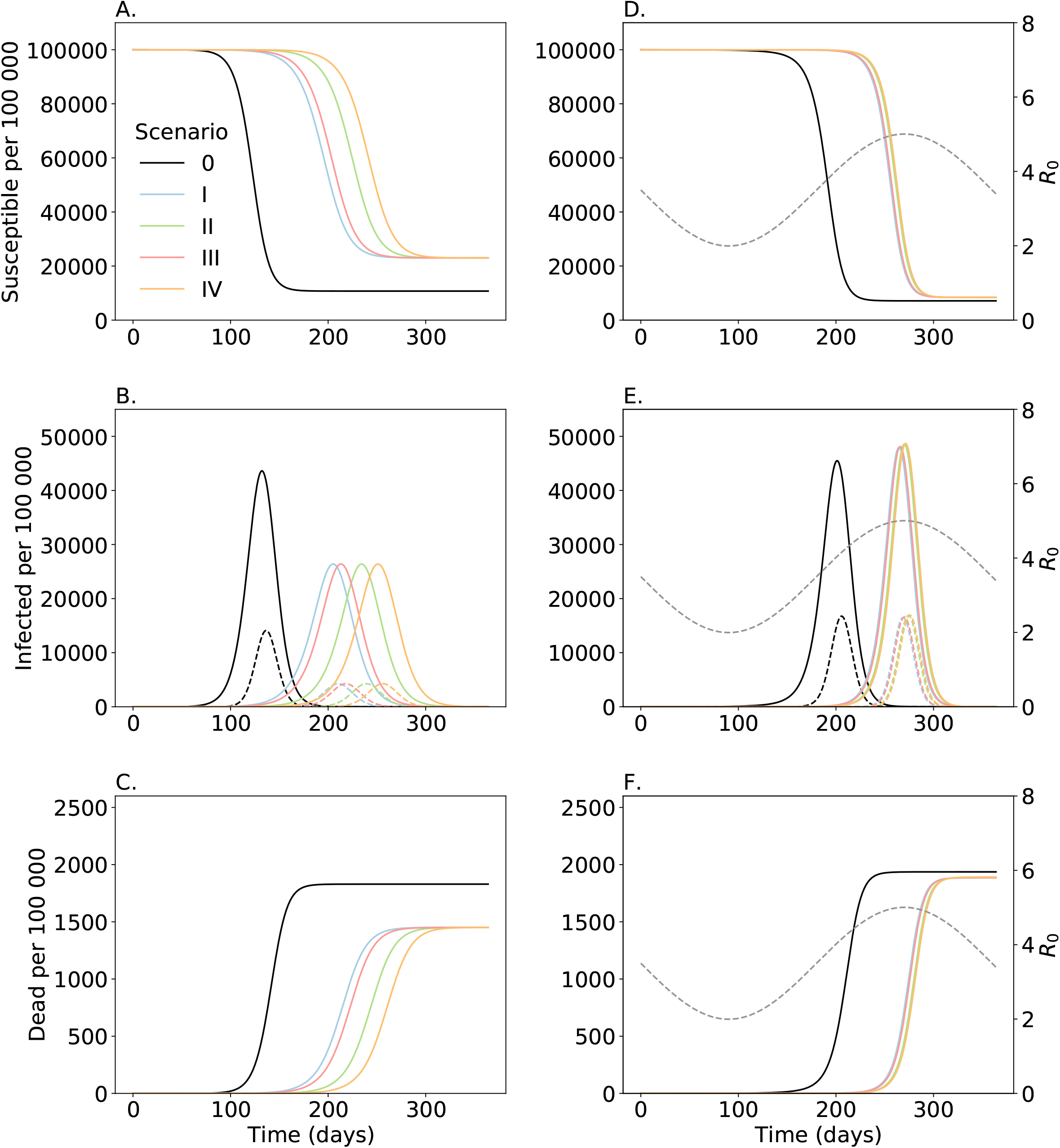
Implication of countermeasures. Shown are the numbers of susceptibles (A, D); (B, E) infected consisting of all latent, prodromal, fully, and late contagious individuals among all single and multi infections (solid lines) and non-transient multi infections (dashed lines); and dead individuals (C, F). Colors are the different scenarios of countermeasures as specified in Table 2. Figs A-C assume no seasonal fluctuations in *R*_0_, while D-F assume seasonal fluctuations in *R*_0_ illustrated by the gray dashed line corresponding to the y-axis on the right-hand side. Parameters are given in S1 Table – S4 Table.

With seasonal fluctuations the epidemic peak falls into the fall period where *R*_0_ starts to increase in the absence of interventions (scenario 0). However, general contact reduction delays the epidemic peak close to the peak of the flu season. This results in a sharper increase in the number of infections and hence a higher epidemic peak. However, it results in slightly less multi infections, because the epidemic peak is narrow and multi infections can only be acquired in substantial numbers in a brief time window (Fig 7D, E). Thus, the overall number of deaths is slightly reduced (Fig 7F).

The increase in infections caused by the first period of general contact reduction (lockdown) can be compensated by a second lockdown as reflected by scenarios (V)-(IX) (Fig 8, S1 Fig). These scenarios correspond to (I) but with a second period of general contact reduction for 45 days with different onsets. Without seasonal fluctuations (reported only for completeness here) the second lockdown has almost no effect as it in occurring too late. This changes with seasonal variation.

**Fig 8.**
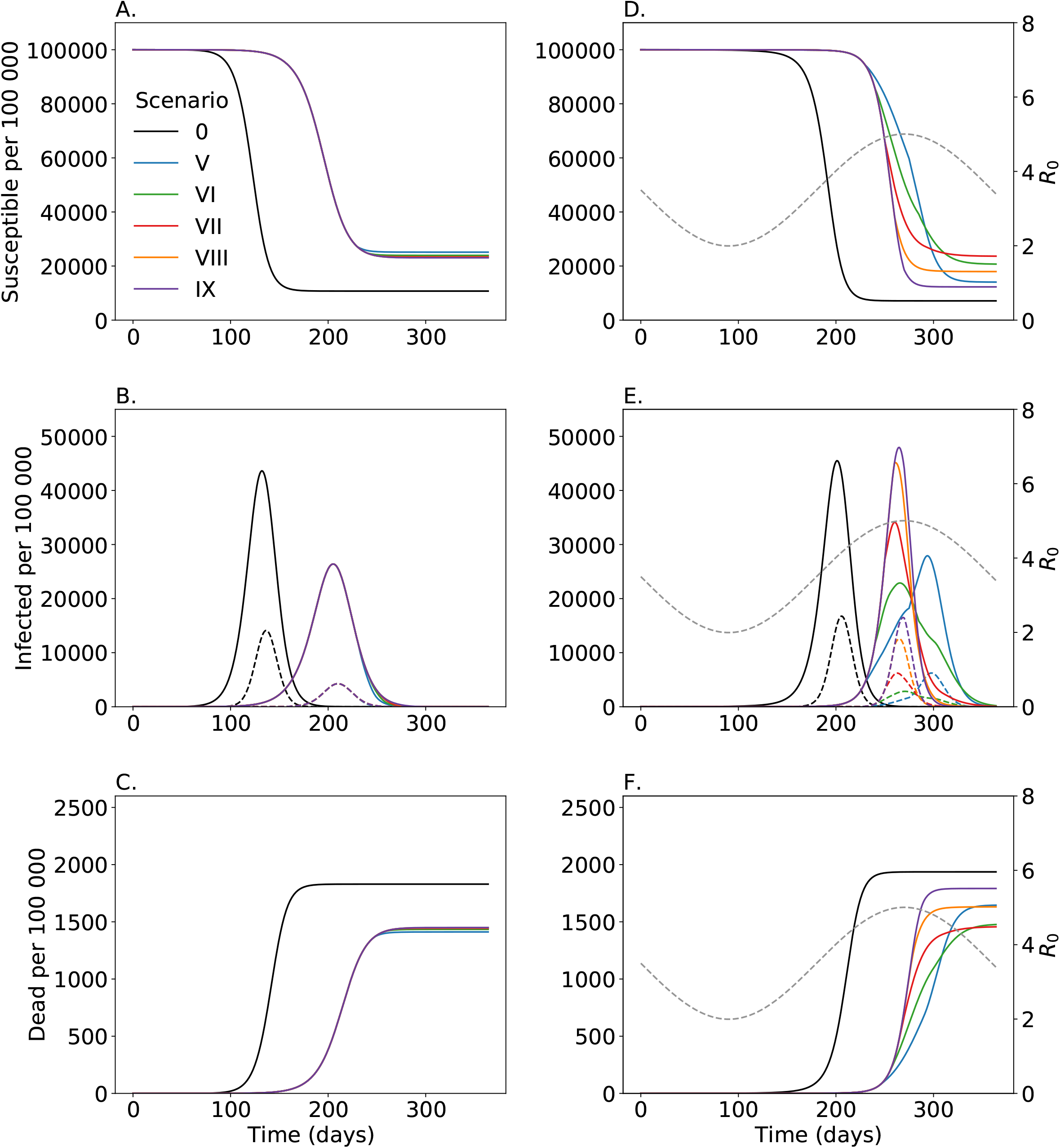
Implication of second lockdown. Shown are the numbers of susceptibles (A, D); (B, E) infected consisting of all latent, prodromal, fully, and late contagious individuals among all single and multi infections (solid lines) and non-transient multi infections (dashed lines); and dead individuals (C, F). Colors are the different scenarios of countermeasures assuming a second lockdown as specified in Table 2. Figs A-C assume no seasonal fluctuations in *R*_0_, while D-F assume seasonal fluctuations in *R*_0_ illustrated by the gray dashed line corresponding to the y-axis on the right-hand side. Parameters are given in S1 Table – S4 Table.

If the second lockdown is properly timed, the epidemic peak is reduced because it is delayed until *R*_0_ decreases (Fig 8E, S1 Fig). The earlier the second lockdown is implemented and falls into the onset of the pandemic peak the more it is delayed. Hence, more time is gained to develop an effective treatment and to prepare for the worst. Moreover, if the lockdown is not implemented too early or too late, the epidemic peak is flattened and widened. Early lockdowns have the advantage that they can be extended to further reduce mortality and the number of cases. On the other side, if lockdowns are implemented too late (and fall in the decline of the epidemic peak), they are inefficient (Fig 8E, S1 Fig).

To highlight the effect of multi infections, the above scenarios can be also studied under the assumption that multi infections do not lead to higher morbidity or mortality, i.e., 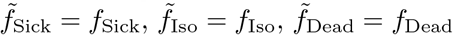. From Fig S2 Fig it is clear that higher morbidity and mortality of multi infections, lead to a slight reduction in the total number of infections occurring during the epidemic (Fig S2 FigA), a slightly lower endemic peak (Fig S2 FigB), but a higher case fatality (Fig S2 FigC) and a substantially higher number of deaths (Fig S2 FigD).

## Discussion

A substantial number of COVID-19 infections was reported among frontline healthcare workers [5, 19, 22]. Particularly, their role in the initial spread of the pandemic was recognized [23]. Partly this is due to improper use of personal protective equipment (PPE) [3, 24]. Frontline healthcare workers are exposed (i) relatively long to infected individuals, and (ii) to multiple infective contacts with different patients within a short time period. Such infections can lead to a higher viral load upon infection or superinfections with different viral variants during the course of the disease. Notably, higher viral loads have been associated with more severe disease [15, 25] – in line with a high number of severe infections among healthcare workers [4]. A similar logic applies to persons in initial epicenters of the global pandemic, e.g., in skiing resorts in northern Italy [26, 27]. Multiple infectious contacts and longer (average) exposure to the virus is initially restricted to certain risk groups, but will become common during the pandemic peak. This is an overseen threat by advocates of herd immunity.

Viral diversity is so high that individuals acquire different viral variants at different infective events. Another overseen risk factor is the increasing viral diversity during an epidemic [cf. 28] – leading to frequent infections with different viral variants when the pandemic peaks.

We studied the potential effect of increased morbidity and mortality due to ‘multi infections’, i.e., infections characterized by a higher viral diversity or higher viral load – acquired either upon infection or during successive infections during one disease episode. For this purpose, we adapted the model underlying the pandemic preparedness tool CovidSim [cf. 20, http://covidsim.eu/], which incorporates general contact reduction during ‘lockdowns’ and case isolation. The model was extended to distinguish between single infections, i.e., infections characterized by less viral diversity and lower viral load upon infection, and multi infections. The latter are characterized by higher morbidity (i.e., more infections are symptomatic, and more symptomatic infections need medical care) and mortality (i.e., more symptomatic infections are lethal). We numerically investigated potential model dynamics reflecting the situation in the East Coast of the USA under a range of parameters summarizing morbidity and mortality of multi infections.

Higher morbidity associated with multi infections leads to a reduction of episodes (if case isolation is sustained) – multi infections are more severe and thus more likely be detected and isolated. However, the reduction in the number of infections has to be interpreted with caution: overall mortality is still higher due to multi infections.

Moreover, more infections are severe leading to an increased demand for medical care and potentially long-term health damages. Estimates of case fatality and disease severity at the onset of a pandemic, when multi infections are rare, will therefore be underestimated. This also includes the estimated demand for medical capacities, treatment costs, and long-term costs due to permanent health damages. Our results show that in the absence of a second lockdown, the epidemic peak will be shifted in the flu season, leading to a higher and narrower peak. This disproportionally increases the number of multi infections and hence deaths. The effects are mediated by (i) the increase in mortality in multi infections, (ii) the likelihood of acquiring a second infection during the various infected phases (before immunity) is reached, and (iii) the increase in the likelihood of developing symptoms. Concluding, increased mortality due to multi infections is a potential threat.

Compared with an epidemic without interventions (reflecting herd immunity), general contact reduction during an initial lockdown and sustained case isolation leads to a slight reduction in the number of deaths, but a substantial delay of the epidemic peak. We therefore also investigated the effects of a second phase of relatively mild contact reduction (lockdown) during the so called ‘second wave’. If it is properly timed, i.e., not too early (when the number of infections is still low), and not too late (when the epidemic peak is already reached), the reduction in the epidemic peak can be achieved as it will be shifted into the fading flu season. This leads to a substantial reduction of the number of cases, multi infections, and mortality. The simulations have to be interpreted with caution. Seasonal fluctuations are unpredictable as well as human behavior during a pandemic. Therefore, our results elucidate qualitative mechanism rather than precise quantitative predictions.

Our results are intended to demonstrate the hypothetical effect of multi infections in simple and straightforward setups. We assume that individuals become permanently immune if they do not die of COVID-19. This neglects temporal immunity [29, 30]. Therefore, the number of superinfections and derived deaths can further increase. Moreover, we did not consider self-imposed distancing measures, i.e., as the number of cases increases, individuals will avoid contacts, have a higher propensity to wear facial masks, etc. This has two reasons: (i) anticipation of human behavior would be speculative and can never be adequately captured by a model; (ii) we wanted to explore the effect in a scenario that aims for herd immunity. Hence, we only wanted to incorporate the most evident control interventions. Nevertheless, it is easy to adapt the model to incorporate more control interventions.

Empirical evidence for multi infections is notoriously difficult to obtain at the beginning of a pandemic. Although the number of COVID-19 infections is threatening, at an early stage, without a complete understanding of the pathogenesis, it is impossible to design a proper study of multi infections. Namely, a representative sample size cannot be achieved despite the many confounding factors.

Notably, multi infections are not the only danger when aiming for herd immunity. An uncontrolled (or hardly controlled) pandemic inevitably renders the virus endemic. This will challenge future SARS-CoV-2 eradication campaigns, as also animals (including, e.g., cats and dogs) become a reservoir for transmission that is difficult to control. Moreover, viral diversity will increase, and aggressive forms of the virus might spread. Viral diversity also challenges vaccination campaigns. Namely, vaccines will need to be adjusted yearly, as it is the case for flu vaccines. Thus, many rounds of vaccinations will be necessary to eradicate the virus. This is particularly dangerous in the context of antibody dependent enhancement, which has been reported in corona viruses, and is thought to be a potential obstacle in vaccination development [31, 32].

Summarizing, increased morbidity and mortality due to multi infections is an important but overseen risk, particularly in the context of herd immunity. Multi infections will become more relevant during the course of the pandemic, when viral diversity and disease prevalence increases. Anyhow, evidence-based research on multi infections is necessary.

## Supporting information

Mathematical Appendix

## Data Availability

Simulated data is used. The source code will be made available upon publication

## Acknowledgments

We want to dedicate this work to all the victims of the SARS-CoV-2 virus. Our grief is with friends and families of the dreadful disease. The authors like to express their sympathy to all working to find a cure for the virus. The authors gratefully acknowledge the helpful comments and discussions with Prof. Martin Eichner on the model.

## Supporting information

**S1 Fig. Implications of a second lockdown**. Shown are the numbers of infected (including latent, prodromal, fully contagious, and late infectious) single and multi infected individuals (solid) and multi infected individuals only (dashed lines). Seasonally vary *R*_0_ is indicated by the gray dashed line corresponding to the y-axis on the right-hand side. Different panels correspond to scenarios described in Table 2. Periods during which case isolation is sustained are indicated in yellow and periods of lockdown are indicated in cyan (overlapping the periods of case isolation). Parameters are given in S1 Table – S4 Table

**S2 Fig. Implications of a second lockdown**. For each scenario (0, V – IX), values are given for the assumption that multi infections cause higher morbidity and mortality and are more likely to be isolated than single infections as described in the text (a) and for the assumption that morbidity, mortality and isolation are the same as for single infection (b). Shown are the total number of deaths (A); (B) case fatality (i.e., deaths per infected individuals); (C) total percentage (of population) being infected or multi infected; and (D) the maximal number of individuals infected or multi infected at the epidemic peak, for scenarios as specified in Table 2 contrasting the case with increased morbidity and mortality of multi infections (a) and no effect of multi infections on mortality and morbidity (b), i.e., the case in which single and multi infections are indistinguishable. The horizontal lines in scenario (a) in (C) and (D) indicate the number of multi infections characterized by increased morbidity and mortality. Parameters are given in S1 Table – S4 Table.

**S1 Appendix. Mathematical Description**

**S1 Table. Population size and compartments**.

**S2 Table. Summary of model parameters**.

**S3 Table. Summary of model parameters**.

**S4 Table. Summary of model parameters**.

