## Supplementary material for "Is increased mortality by multiple exposures to COVID-19 an overseen factor when aiming for herd immunity?": Mathematical Appendix

### Appendix: Mathematical description

We study the hypothetical effect of multiple infections with COVID-19 during one infective period on disease severity and mortality by an extended SEIR model (see Figure 1 for an illustration). Particularly, we adapt the deterministic compartmental model underlying the pandemic preparedness tool CovidSIM [1].

#### Model compartments

A population of $N$ individuals is assumed. The model follows the time change in the number of individuals being susceptible ($S$), latently infected ($E_{\text{Sum}}$) consisting of $n_E$ sub-states ($E_k$, $k = 1, \dots, n_E$), prodromal ($P_{\text{Sum}}$) consisting of $n_P$ sub-states ($P_k$, $k = 1, \dots, n_P$), fully contagious ($I_{\text{Sum}}$) consisting of $n_I$ sub-states ($I_k$, $k = 1, \dots, n_I$), late infectious ($L_{\text{Sum}}$) consisting of $n_L$ sub-states ($L_k$, $k = 1, \dots, n_L$), in the final "removed" stage ($R$), and dead individuals ($D$).

A fraction $f_{\text{Sick}}$ of fully contagious individuals becomes symptomatic, i.e., they get sick, while the remaining infections are asymptomatic. Asymptomatic infections are never lethal, whereas a fraction $f_{\text{Dead}}$ of symptomatic infections results in death. If individuals acquire multi-infections they are more likely to become symptomatic (denoted $\tilde{f}_{\text{Sick}}$) and die (denoted $\tilde{f}_{\text{Dead}}$). Therefore, we have to distinguish between single- and multi-infected individuals. A multi-infection can be acquired at one infective contact with a multi-infected individual or by successive infections. In the latter case, the multi-infection is acquired on top of a single infection and the increased risks of developing symptoms and of mortality do not immediately manifest. Namely, it takes an average duration $1/\alpha$ until the multi-infection becomes effective. This reflects a transient phase during which neither the likelihoods of developing symptoms nor the mortality are yet elevated. As a consequence, in all stages of the infection, we need to distinguish between (i) single infections, (ii) multi infections in the transient phase, and (iii) the remaining multi infections. This is done by modeling the three classes by different compartments. The notation of the last paragraph is used for single infections. The respective compartments in multi-infected individuals in the transient phase are denoted by $E_k^*$ ($k = 1, \dots, n_E$), $P_k^*$ ($k = 1, \dots, n_P$), $I_k^*$ ($k = 1, \dots, n_I$), and $L_k^*$ ($k = 1, \dots, n_L$). The compartment of the remaining multi-infected individuals are denoted by $\tilde{E}_k$ ($k = 1, \dots, n_E$), $\tilde{P}_k$ ($k = 1, \dots, n_P$), $\tilde{I}_k$ ($k = 1, \dots, n_I$), and $\tilde{L}_k$ ($k = 1, \dots, n_L$).

##### Total numbers of infected and symptomatic individuals

The total number of single infections in the latent stage is given by

$$E_{\text{Sum}}(t) = \sum_{k=1}^{n_E} E_k(t), \tag{1a}$$

$$\tag{1b}$$

while that of latent multi-infections in the transient phase is

$$E^*_{\text{Sum}}(t) = \sum_{k=1}^{n_E} E^*_k(t), \tag{1c}$$

and that of the remaining multi-infections in the latent stage is

$$\tilde{E}_{\text{Sum}}(t) = \sum_{k=1}^{n_E} \tilde{E}_k(t). \tag{1d}$$

Likewise the number of prodromal single, transient, and remaining multi-infects are

$$P_{\text{Sum}}(t) = \sum_{k=1}^{n_P} P_k(t), \tag{2a}$$

$$P^*_{\text{Sum}}(t) = \sum_{k=1}^{n_P} P^*_k(t), \tag{2b}$$

and

$$\tilde{P}_{\text{Sum}}(t) = \sum_{k=1}^{n_P} \tilde{P}_k(t). \tag{2c}$$

The number of the fully contagious and late infectious individuals in the three classes of infections are, respectively,

$$I_{\text{Sum}}(t) = \sum_{k=1}^{n_I} I_k(t), \tag{3a}$$

$$I^*_{\text{Sum}}(t) = \sum_{k=1}^{n_I} I^*_k(t), \tag{3b}$$

$$\tilde{I}_{\text{Sum}}(t) = \sum_{k=1}^{n_I} \tilde{I}_k(t), \tag{3c}$$

$$L_{\text{Sum}}(t) = \sum_{k=1}^{n_L} L_k(t), \tag{4a}$$

$$L^*_{\text{Sum}}(t) = \sum_{k=1}^{n_L} L^*_k(t), \tag{4b}$$

and

$$\tilde{L}_{\text{Sum}}(t) = \sum_{k=1}^{n_L} \tilde{L}_k(t). \tag{4c}$$

The number of symptomatic infections in the fully contagious phase that are symptomatic among single infections is

$$I_{\text{Sick}}(t) = f_{\text{Sick}} I_{\text{Sum}}(t). \tag{5a}$$

Likewise, the number of symptomatic multi-infections in the fully contagious phase that are transient is

$$I^*_{\text{Sick}}(t) = f_{\text{Sick}} I^*_{\text{Sum}}(t)). \tag{5b}$$

The remaining multi infections have elevated risk of developing symptoms, so that the number of remaining symptomatic multi-infections in the fully contagious phase is

$$\tilde{I}_{\text{Sick}}(t) = \tilde{f}_{\text{Sick}} \tilde{I}_{\text{Sum}}(t). \tag{5c}$$

Similarly the numbers of symptomatic infections in the late infectious phase for the three classes are,

$$L_{\text{Sick}}(t) = f_{\text{Sick}} L_{\text{Sum}}(t), \tag{6a}$$
$$L^*_{\text{Sick}}(t) = f_{\text{Sick}} L^*_{\text{Sum}}(t), \tag{6b}$$

and

$$\tilde{L}_{\text{Sick}}(t) = \tilde{f}_{\text{Sick}} \tilde{L}_{\text{Sum}}(t), \tag{6c}$$

respectively.

#### Case Isolation

A fraction $f_{\text{Iso}}$ of symptomatic single and multi-infections in the transient phase get hospitalized and will be put into quarantine wards. Moreover, a (higher) fraction $\tilde{f}_{\text{Iso}}$ of the remaining symptomatic multi infections are isolated, reflecting that these infections lead to more severe symptoms and hence a higher demand for medical help. Infected individuals that cannot be isolated in quarantine wards because their maximum capacity ($Q_{\text{max}}$) is reached are sent into home isolation. While isolation in wards is perfect, in home isolation only a fraction $p_{\text{Home}}$ of infectious contacts is prevented. Isolation lasts until recovery or death. Importantly, case isolation mechanisms are sustained only during the time interval from $t_{\text{Iso}_1}$ to $t_{\text{Iso}_2}$. In particular, case isolation policies might not be in place at the introduction of the disease. At time $t$ ($t_{\text{Iso}_1} \leq t \leq t_{\text{Iso}_2}$), the number of individuals isolated in quarantine wards or at home is

$$Q(t) = f_{\text{Iso}} f_{\text{Sick}} \Big( I_{\text{Sum}}(t) + I^*_{\text{Sum}}(t) + L_{\text{Sum}}(t) + L^*_{\text{Sum}}(t) \Big) + \tilde{f}_{\text{Iso}} \tilde{f}_{\text{Sick}} \Big( \tilde{I}_{\text{Sum}}(t) + \tilde{L}_{\text{Sum}}(t) \Big). \tag{7}$$

The capacity of quarantine wards is divided proportionally by individuals in the respective sub-states of the infection. Thus, the number of fully contagious single infections in the $k$th sub-state is

$$I_k^{(\text{Iso})}(t) = \begin{cases} f_{\text{Sick}} f_{\text{Iso}} I_k(t) & \text{if} \quad t_{\text{Iso}_1} \leq t \leq t_{\text{Iso}_2} \quad \text{and} \quad Q(t) \leq Q_{\text{max}}, \\ f_{\text{Sick}} f_{\text{Iso}} I_k(t) \frac{Q_{\text{max}}}{Q(t)} & \text{if} \quad t_{\text{Iso}_1} \leq t \leq t_{\text{Iso}_2} \quad \text{and} \quad Q(t) > Q_{\text{max}}, \\ 0 & \text{else.} \end{cases} \tag{8a}$$

The number of multi infected fully contagious individuals that are in the $k$th sub-state of the transient phase is

$$I_k^{(*,\text{Iso})}(t) = \begin{cases} f_{\text{Sick}} f_{\text{Iso}} I^*_k(t) & \text{if} \quad t_{\text{Iso}_1} \leq t \leq t_{\text{Iso}_2} \quad \text{and} \quad Q(t) \leq Q_{\text{max}}, \\ f_{\text{Sick}} f_{\text{Iso}} I^*_k(t) \frac{Q_{\text{max}}}{Q(t)} & \text{if} \quad t_{\text{Iso}_1} \leq t \leq t_{\text{Iso}_2} \quad \text{and} \quad Q(t) > Q_{\text{max}}, \\ 0 & \text{else,} \end{cases} \tag{8b}$$

whereas the remaining fully contagious multi infections in the $k$ sub-state occupying the quarantine wards is

$$\tilde{I}_k^{(\text{Iso})}(t) = \begin{cases} \tilde{f}_{\text{Sick}} \tilde{f}_{\text{Iso}} \tilde{I}_k(t) & \text{if} \quad t_{\text{Iso}_1} \leq t \leq t_{\text{Iso}_2} \quad \text{and} \quad Q(t) \leq Q_{\text{max}}, \\ \tilde{f}_{\text{Sick}} \tilde{f}_{\text{Iso}} \tilde{I}_k(t) \frac{Q_{\text{max}}}{Q(t)} & \text{if} \quad t_{\text{Iso}_1} \leq t \leq t_{\text{Iso}_2} \quad \text{and} \quad Q(t) > Q_{\text{max}}, \\ 0 & \text{else.} \end{cases} \tag{8c}$$

Similarly, the number of late infectious individuals in the $k$th sub-state in the respective classes of infections are

$$L_k^{(\mathrm{Iso})}(t) = \begin{cases} f_{\mathrm{Sick}} f_{\mathrm{Iso}} L_k(t) & \text{if} \quad t_{\mathrm{Iso}_1} \le t \le t_{\mathrm{Iso}_2} \quad \text{and} \quad Q(t) \le Q_{\max}, \\ f_{\mathrm{Sick}} f_{\mathrm{Iso}} L_k(t) \frac{Q_{\max}}{Q(t)} & \text{if} \quad t_{\mathrm{Iso}_1} \le t \le t_{\mathrm{Iso}_2} \quad \text{and} \quad Q(t) > Q_{\max}, \\ 0 & \text{else}, \end{cases} \tag{9a}$$

$$L_k^{(*,\mathrm{Iso})}(t) = \begin{cases} f_{\mathrm{Sick}} f_{\mathrm{Iso}} L_k^*(t) & \text{if} \quad t_{\mathrm{Iso}_1} \le t \le t_{\mathrm{Iso}_2} \quad \text{and} \quad Q(t) \le Q_{\max}, \\ f_{\mathrm{Sick}} f_{\mathrm{Iso}} L_k^*(t) \frac{Q_{\max}}{Q(t)} & \text{if} \quad t_{\mathrm{Iso}_1} \le t \le t_{\mathrm{Iso}_2} \quad \text{and} \quad Q(t) > Q_{\max}, \\ 0 & \text{else}, \end{cases} \tag{9b}$$

and

$$\tilde{L}_k^{(\mathrm{Iso})}(t) = \begin{cases} \tilde{f}_{\mathrm{Sick}} \tilde{f}_{\mathrm{Iso}} \tilde{L}_k(t) & \text{if} \quad t_{\mathrm{Iso}_1} \le t \le t_{\mathrm{Iso}_2} \quad \text{and} \quad Q(t) \le Q_{\max}, \\ \tilde{f}_{\mathrm{Sick}} \tilde{f}_{\mathrm{Iso}} \tilde{L}_k(t) \frac{Q_{\max}}{Q(t)} & \text{if} \quad t_{\mathrm{Iso}_1} \le t \le t_{\mathrm{Iso}_2} \quad \text{and} \quad Q(t) > Q_{\max}, \\ 0 & \text{else}. \end{cases} \tag{9c}$$

Likewise, the numbers of fully contagious and late infectious single and multiple infections in home isolation are, respectively,

(10a)

$$I_k^{(\mathrm{Home})}(t) = \begin{cases} f_{\mathrm{Sick}} f_{\mathrm{Iso}} I_k(t) \left(1 - \frac{Q_{\max}}{Q(t)}\right) & \text{if } t_{\mathrm{Iso}_1} \le t \le t_{\mathrm{Iso}_2} \quad \text{and} \quad Q(t) > Q_{\max}, \\ 0 & \text{else}, \end{cases}$$

(10b)

$$I_k^{(*,\mathrm{Home})}(t) = \begin{cases} f_{\mathrm{Sick}} f_{\mathrm{Iso}} I_k^*(t) \left(1 - \frac{Q_{\max}}{Q(t)}\right) & \text{if } t_{\mathrm{Iso}_1} \le t \le t_{\mathrm{Iso}_2} \quad \text{and} \quad Q(t) > Q_{\max}, \\ 0 & \text{else}, \end{cases}$$

(10c)

$$\tilde{I}_k^{(\mathrm{Home})}(t) = \begin{cases} \tilde{f}_{\mathrm{Sick}} \tilde{f}_{\mathrm{Iso}} \tilde{I}_k(t) \left(1 - \frac{Q_{\max}}{Q(t)}\right) & \text{if } t_{\mathrm{Iso}_1} \le t \le t_{\mathrm{Iso}_2} \quad \text{and} \quad Q(t) > Q_{\max}, \\ 0 & \text{else}, \end{cases}$$

(10d)

and

$$L_k^{(\mathrm{Home})}(t) = \begin{cases} f_{\mathrm{Sick}} f_{\mathrm{Iso}} L_k(t) \left(1 - \frac{Q_{\max}}{Q(t)}\right) & \text{if } t_{\mathrm{Iso}_1} \le t \le t_{\mathrm{Iso}_2} \quad \text{and} \quad Q(t) > Q_{\max}, \\ 0 & \text{else}, \end{cases}$$

(11a)

$$L_k^{(*,\mathrm{Home})}(t) = \begin{cases} f_{\mathrm{Sick}} f_{\mathrm{Iso}} L_k^*(t) \left(1 - \frac{Q_{\max}}{Q(t)}\right) & \text{if } t_{\mathrm{Iso}_1} \le t \le t_{\mathrm{Iso}_2} \quad \text{and} \quad Q(t) > Q_{\max}, \\ 0 & \text{else}, \end{cases}$$

(11b)

$$\tilde{L}_k^{(\mathrm{Home})}(t) = \begin{cases} \tilde{f}_{\mathrm{Sick}} \tilde{f}_{\mathrm{Iso}} \tilde{L}_k(t) \left(1 - \frac{Q_{\max}}{Q(t)}\right) & \text{if } t_{\mathrm{Iso}_1} \le t \le t_{\mathrm{Iso}_2} \quad \text{and} \quad Q(t) > Q_{\max}, \\ 0 & \text{else}. \end{cases}$$

(11c)

Because infectious contacts are avoided by isolation measures, not all infected individuals participate in transmission. The effective number of fully contagious single infections in the $k$th sub-state that effectively participate in disease transmission at time $t$ is

$$I_k^{(\mathrm{Eff})}(t) = I_k(t) - I_k^{(\mathrm{Iso})}(t) - p_{\mathrm{Home}} I_k^{(\mathrm{Home})}(t). \tag{12a}$$

The effective number of multi-infected fully contagious individuals in the transient phase in the $k$th sub-state is

$$I_k^{(*,\mathrm{Eff})}(t) = I_k^*(t) - I_k^{(*,\mathrm{Iso})}(t) - p_{\mathrm{Home}} I_k^{(*,\mathrm{Home})}(t). \tag{12b}$$

Finally, the remaining multi-infected fully contagious individuals in the $k$th sub-state participating in infections is

$$\tilde{I}_k^{(\mathrm{Eff})}(t) = \tilde{I}_k(t) - \tilde{I}_k^{(\mathrm{Iso})}(t) - p_{\mathrm{Home}} \tilde{I}_k^{(\mathrm{Home})}(t). \tag{12c}$$

This results in the total numbers of fully contagious individuals participating in infection in the three classes given by

$$I_{\mathrm{Eff}}(t) = \sum_{k=1}^{n_I} I_k^{(\mathrm{Eff})}(t), \tag{13a}$$

$$I_{\mathrm{Eff}}^*(t) = \sum_{k=1}^{n_I} I_k^{(*,\mathrm{Eff})}(t), \tag{13b}$$

and

$$\tilde{I}_{\mathrm{Eff}}(t) = \sum_{k=1}^{n_I} \tilde{I}_k^{(\mathrm{Eff})}(t). \tag{13c}$$

As for the fully contagious individuals the effective numbers of individuals in the $k$th late infectious sub-states contributing to transmission in the three classes are

$$L_k^{(\mathrm{Eff})}(t) = L_k(t) - L_k^{(\mathrm{Iso})}(t) - p_{\mathrm{Home}} L_k^{(\mathrm{Home})}(t), \tag{14a}$$

$$L_k^{(*,\mathrm{Eff})}(t) = L_k^*(t) - L_k^{(*,\mathrm{Iso})}(t) - p_{\mathrm{Home}} L_k^{(*,\mathrm{Home})}(t), \tag{14b}$$

and

$$\tilde{L}_k^{(\mathrm{Eff})}(t) = \tilde{L}_k(t) - \tilde{L}_k^{(\mathrm{Iso})}(t) - p_{\mathrm{Home}} \tilde{L}_k^{(\mathrm{Home})}(t). \tag{14c}$$

Likewise the effective numbers of late infectious individuals in the three classes that participate in infection are

$$L_{\mathrm{Eff}}(t) = \sum_{k=1}^{n_L} L_k^{(\mathrm{Eff})}(t), \tag{15a}$$

$$L_{\mathrm{Eff}}^*(t) = \sum_{k=1}^{n_L} L_k^{(*,\mathrm{Eff})}(t), \tag{15b}$$

and

$$\tilde{L}_{\mathrm{Eff}}(t) = \sum_{k=1}^{n_L} \tilde{L}_k^{(\mathrm{Eff})}(t), \tag{15c}$$

#### Durations

The average duration of the latent, prodromal, fully contagious and late infectious periods are denoted by $D_E$, $D_P$, $D_I$, and $D_L$, respectively. Infected individuals have to progress through $n_E$ equivalent latent sub-states. Hence, the average duration in each latent sub-state is $D_E/n_E$, and individuals leave the latent sub-states at a rate

$$\varepsilon := \frac{n_E}{D_E}. \tag{16a}$$

As a consequence the duration in the latent phase is Erlang distributed with mean $D_E$ and variance $D_E/n_E$. Similarly, the rates at which individuals leave the prodromal, fully contagious, and late infectious states are, respectively,

$$\varphi := \frac{n_P}{D_P}, \quad \gamma := \frac{n_I}{D_I}, \quad \text{and} \quad \delta := \frac{n_L}{D_L}. \tag{16b}$$

As outlined above, if a single infected individual is infected again during the latent, prodromal, fully contagious, or late infectious phases, it becomes multi infected, but the multi infection does not manifest immediately. Namely, it the multi-infection is transient for an average duration $D_T = 1/\alpha$.

#### Contact rates

The basic reproduction number $R_0$ is the average number of infections caused by an infected individual during the whole duration of the infections consisting of the latent, prodromal, fully contagious and late infectious periods. The quantity $R_0$ fluctuates seasonally with an amplitude $a$ and a peak at time $t_{R_{0\max}}$, modeled by

$$R_0(t) = \bar{R}_0\left(1 + a\cos\left(2\pi\frac{t - t_{R_{0\max}}}{365}\right)\right). \tag{17}$$

Susceptibles encounter infected individuals (not in isolation) randomly. The relative contagiousness of prodromal and late infectious individuals compared with fully contagious ones are $c_P$ and $c_L$, respectively. Single and multi-infected individuals are equally contagious in the respective stages. Thus, the effective contact rates are

$$\beta_P(t) = \frac{c_P\bar{R}_0(t)}{c_P D_P + D_I + c_L D_L}, \tag{18}$$

$$\beta_I(t) = \frac{\bar{R}_0(t)}{c_P D_P + D_I + c_L D_L}, \tag{19}$$

$$\beta_L(t) = \frac{c_L\bar{R}_0(t)}{c_P D_P + D_I + c_L D_L}. \tag{20}$$

#### General contact reduction and force of infection

During the time interval from $t_{\text{Dist}_1}$, $t_{\text{Dist}_2}$, general contact recusing measures, e.g., social distancing, curfews, etc., are in place, which reduce contacts by an amount $p_{\text{Dist}}$. We define

$$p_{\text{Gen}}(t) = \begin{cases} p_{\text{Dist}} & \text{for} \quad t_{\text{Dist}_1} \le t \le t_{\text{Dist}_2}, \\ 0 & \text{else.} \end{cases} \tag{21}$$

An infectious contact with a multi-infected individual leads to a multi infection with probability $\tilde{m}$. Thus, the force of infection by which susceptible acquire single infections is given by

$$\lambda^{(s)}(t) = \lambda^{(s)}_{\text{Ext}} + \big(1 - p_{\text{Gen}}(t)\big)\Bigg(\beta_P(t)\Big(P_{\text{Sum}}(t) + P^*_{\text{Sum}}(t) + (1-\tilde{m})\tilde{P}_{\text{Sum}}(t)\Big) + \beta_I(t)\Big(I_{\text{Eff}}(t) + I^*_{\text{Eff}}(t) + (1-\tilde{m})\tilde{I}_{\text{Eff}}(t)\Big) + \beta_L(t)\Big(L_{\text{Eff}}(t) + L^*_{\text{Eff}}(t) + (1-\tilde{m})\tilde{L}_{\text{Eff}}(t)\Big)\Bigg), \tag{22}$$

where $\lambda^{(s)}_{\text{Ext}}$ is the external force of infection leading to single infections form outside the population. Likewise the force of infections by which susceptibles acquire multi infections is given by

$$\lambda^{(m)}(t) = \lambda^{(m)}_{\text{Ext}} + \big(1 - p_{\text{Gen}}(t)\big)\Big(\beta_P(t)\tilde{m}\tilde{P}_{\text{Sum}}(t) + \beta_I(t)\tilde{m}\tilde{I}_{\text{Eff}}(t) + \beta_L(t)\tilde{m}\tilde{L}_{\text{Eff}}(t)\Big), \tag{23}$$

where $\lambda^{(m)}_{\text{Ext}}$ is the external force of infection leading to multi infections form outside the population. The combined force of infections by which susceptibles become infected is

$$\lambda(t) = \lambda^{(s)}(t) + \lambda^{(m)}(t). \tag{24}$$

Single infected individuals at any state of the infection can acquire a multi infection. However, the initial infection also leads to some protection as the immune system is already activated. Thus the force of infection experienced by latent, prodromal, fully infected, and late infectious individuals is reduced by factors $q_E$, $q_P$, $q_I$, $q_L$, respectively.

#### Differential Equation

Putting together what is described above leads to the following system of differential equations (cf. Fig 1). The change in the number of susceptible individuals is given by

$$\frac{dS(t)}{dt} = -\lambda(t)\frac{S(t)}{N}. \tag{25}$$

The number of single infected individuals in the latent sub-states change according to

$$\frac{dE_1(t)}{dt} = \lambda^{(s)}(t)\frac{S(t)}{N} - \varepsilon E_1(t) - q_E\lambda(t)\frac{E_1(t)}{N}, \tag{26a}$$

$$\frac{dE_k(t)}{dt} = \varepsilon E_{k-1}(t) - \varepsilon E_k(t) - q_E\lambda(t)\frac{E_k(t)}{N} \quad \text{for} \quad 2 \le k \le n_E. \tag{26b}$$

The dynamics of transient multi infections in the latent phase are

$$\frac{dE^*_1(t)}{dt} = q_E\lambda(t)\frac{E_1(t)}{N} - \varepsilon E^*_1(t) - \alpha E^*_1(t), \tag{26c}$$

$$\frac{dE^*_k(t)}{dt} = q_E\lambda(t)\frac{E_k(t)}{N} + \varepsilon E^*_{k-1}(t) - \varepsilon E^*_k(t) - \alpha E^*_k(t) \quad \text{for} \quad 2 \le k \le n_E, \tag{26d}$$

whereas the numbers of reaming multi-infected individuals in the latent phase change according to

$$\frac{d\tilde{E}_1(t)}{dt} = \lambda^{(m)}(t)\frac{S(t)}{N} + \alpha E^*_1(t) - \varepsilon\tilde{E}_1(t), \tag{26e}$$

$$\frac{d\tilde{E}_k(t)}{dt} = \alpha E^*_k(t) + \varepsilon\tilde{E}_{k-1}(t) - \varepsilon\tilde{E}_k(t) \quad \text{for} \quad 2 \le k \le n_E. \tag{26f}$$

Likewise the number of prodromal individuals in the various classes of infections and sub-states change according to

$$\frac{dP_1(t)}{dt} = \varepsilon E_{n_E}(t) - \varphi P_1(t) - q_P \lambda(t) \frac{P_1(t)}{N}, \tag{27a}$$

$$\frac{dP_k(t)}{dt} = \varphi P_{k-1}(t) - \varphi P_k(t) - q_P \lambda(t) \frac{P_k(t)}{N} \qquad \text{for} \quad 2 \leq k \leq n_P, \tag{27b}$$

$$\frac{dP_1^*(t)}{dt} = q_P \lambda(t) \frac{P_1(t)}{N} + \varepsilon E_{n_E}^*(t) - \varphi P_1^*(t) - \alpha P_1^*(t), \tag{27c}$$

$$\frac{dP_k^*(t)}{dt} = q_P \lambda(t) \frac{P_k(t)}{N} + \varphi P_{k-1}^*(t) - \varphi P_k^*(t) - \alpha P_k^*(t) \qquad \text{for} \quad 2 \leq k \leq n_P, \tag{27d}$$

$$\frac{d\tilde{P}_1(t)}{dt} = \alpha P_1^*(t) + \varepsilon \tilde{E}_{n_E}(t) - \varphi \tilde{P}_1(t), \tag{27e}$$

$$\frac{d\tilde{P}_k(t)}{dt} = \alpha P_k^*(t) + \varphi \tilde{P}_{k-1}(t) - \varphi \tilde{P}_k(t) \qquad \text{for} \quad 2 \leq k \leq n_P. \tag{27f}$$

For the number of fully contagious individuals the differential equations become

$$\frac{dI_1(t)}{dt} = \varphi P_{n_P}(t) - \gamma I_1(t) - q_I \lambda(t) \frac{I_1^{(\mathrm{Eff})}(t)}{N}, \tag{28a}$$

$$\frac{dI_k(t)}{dt} = \gamma I_{k-1}(t) - \gamma P_k(t) - q_I \lambda(t) \frac{I_k^{(\mathrm{Eff})}(t)}{N} \qquad \text{for} \quad 2 \leq k \leq n_I, \tag{28b}$$

$$\frac{dI_1^*(t)}{dt} = q_I \lambda(t) \frac{I_1^{(\mathrm{Eff})}(t)}{N} + \varphi P_{n_P}^*(t) - \gamma I_1^*(t) - \alpha I_1^*(t), \tag{28c}$$

$$\frac{dI_k^*(t)}{dt} = q_I \lambda(t) \frac{I_k^{(\mathrm{Eff})}(t)}{N} + \gamma I_{k-1}^*(t) - \gamma I_k^*(t) - \alpha I_k^*(t) \qquad \text{for} \quad 2 \leq k \leq n_I, \tag{28d}$$

$$\frac{d\tilde{I}_1(t)}{dt} = \alpha I_1^*(t) + \varphi \tilde{P}_{n_P}(t) - \gamma \tilde{I}_1(t), \tag{28e}$$

$$\frac{d\tilde{I}_k(t)}{dt} = \alpha I_k^*(t) + \gamma \tilde{I}_{k-1}(t) - \gamma \tilde{I}_k(t) \qquad \text{for} \quad 2 \leq k \leq n_I, \tag{28f}$$

The number of late infectious individuals change according to

$$\frac{dL_1(t)}{dt} = \gamma I_{n_I}(t) - \delta L_1(t) - q_L \lambda(t) \frac{L_1^{(\mathrm{Eff})}(t)}{N}, \tag{29a}$$

$$\frac{dL_k(t)}{dt} = \delta L_{k-1}(t) - \delta L_k(t) - q_L \lambda(t) \frac{L_k^{(\mathrm{Eff})}(t)}{N} \qquad \text{for} \quad 2 \leq k \leq n_L, \tag{29b}$$

$$\frac{dL_1^*(t)}{dt} = q_L \lambda(t) \frac{L_1^{(\mathrm{Eff})}(t)}{N} + \gamma I_{n_I}^*(t) - \delta L_1^*(t) - \alpha L_1^*(t), \tag{29c}$$

$$\frac{dL_k^*(t)}{dt} = q_L \lambda(t) \frac{L_k^{(\mathrm{Eff})}(t)}{N} + \delta L_{k-1}^*(t) - \delta L_k^*(t) - \alpha L_k^*(t) \qquad \text{for} \quad 2 \leq k \leq n_L, \tag{29d}$$

$$\frac{d\tilde{L}_1(t)}{dt} = \alpha L_1^*(t) + \gamma \tilde{I}_{n_I}(t) - \delta \tilde{L}_1(t), \tag{29e}$$

$$\frac{d\tilde{L}_k(t)}{dt} = \alpha L_k^*(t) + \delta \tilde{L}_{k-1}(t) - \delta \tilde{L}_k(t) \qquad \text{for} \quad 2 \leq k \leq n_L. \tag{29f}$$

Finally, the number of recovered and dead individuals change according to

$$\frac{dR(t)}{dt} = \delta\big(1 - f_{\mathrm{Sick}} f_{\mathrm{Dead}}\big)\big(L_{n_L}(t) + L_{n_L}^*(t)\big) + \delta\big(1 - \tilde{f}_{\mathrm{Sick}} \tilde{f}_{\mathrm{Dead}}\big)\tilde{L}_{n_L}(t), \tag{30}$$

and

$$\frac{dD(t)}{dt} = \delta f_{\text{Sick}} f_{\text{Dead}}\big(L_{n_L}(t) + L^*_{n_L}(t)\big) + \delta \tilde{f}_{\text{Sick}} \tilde{f}_{\text{Dead}} \tilde{L}_{n_L}(t). \tag{31}$$

#### Number of multi infections

The cumulative number of multi infections at time $T$, excluding those, that were still in the transient phase upon recovery or death, can be calculated as follows. The number of single and multi infections at time $T$ is $N - S(T)$, while the number of deaths that occurred until then is $D(T)$. Denoting the proportion of deaths attributed to multi infections (excluding those that were transient upon recovery) by $x$, we have the equation

$$D(T) = \Big(N - S(T)\Big)\big(x\tilde{f}_{\text{Sick}}\tilde{f}_{\text{Dead}} + (1-x) f_{\text{Sick}} f_{\text{Dead}}\big), \tag{32}$$

yielding the total number of multi infections (excluding those that were transient upon recovery and death) as

$$\Big(N - S(T)\Big)x = \Big(N - S(T)\Big)\frac{D(T) - f_{\text{Sick}} f_{\text{Dead}}\big(N - S(T)\big)}{\tilde{f}_{\text{Sick}}\tilde{f}_{\text{Dead}} - f_{\text{Sick}} f_{\text{Dead}}}. \tag{33}$$
