## Supplementary material for "Is increased mortality by multiple exposures to COVID-19 an overseen factor when aiming for herd immunity?": Table S1

**S1 Table.** Population size and compartments.

| Parameter | Description | value |
|---|---|---|
| $N$ | Population size | 250,000,000 |
| $S(0)$ | No. susceptible | 249,999,999 |
| $E_k(0)$ | No. single infected in $k$th latent states ( $1 \leq k \leq n_E$) | 0 |
| $E_k^*(0)$ | No. transient multi infections in latent states ($1 \leq k \leq n_E$) | 0 |
| $\tilde{E}_k(0)$ | No. multi infected in $k$th latent states ($1 \leq k \leq n_E$) | 0 |
| $P_k(0)$ | No. single infected in $k$th prodromal states ( $1 \leq k \leq n_P$) | 0 |
| $P_k^*(0)$ | No. transient multi infections in prodromal states ($1 \leq k \leq n_P$) | 0 |
| $\tilde{P}_k(0)$ | No. multi infected in prodromal states ( $1 \leq k \leq n_P$) | 0 |
| $I_1(0)$ | No. single infected in first fully contagious Erlang state | 1 |
| $I_k(0)$ | No. single infected in $k$th fully contagious states ($2 \leq k \leq n_I$) | 0 |
| $I_k^*(0)$ | No. transient multi infections in $k$th fully contagious states ($1 \leq k \leq n_I$) | 0 |
| $\tilde{I}_k(0)$ | No. multi infected in full contagious states ($1 \leq k \leq n_I$ ) | 0 |
| $L_k(0)$ | No. single infected in $k$th late-infectious states ($1 \leq k \leq n_L$ ) | 0 |
| $L_k^*(0)$ | No. transient multi infections in $k$th late-infectious states ($1 \leq k \leq n_L$) | 0 |
| $\tilde{L}_k(0)$ | No. multi infected in $k$th late-infectious states ($1 \leq k \leq n_L$) | 0 |
| $R$ | No. recovered | 0 |
| $D$ | No. dead | 0 |

Population size and compartments and their respective parameter choices.
