## Supplementary material for "Is increased mortality by multiple exposures to COVID-19 an overseen factor when aiming for herd immunity?": Table S2

**S2 Table.** Summary of model parameters.

| Parameter | Definition | Value |
|---|---|---|
| $n_{\mathrm{E}}$ | No. of latency Erlang states | 16 |
| $n_{\mathrm{P}}$ | No. of prodromal Erlang states | 16 |
| $n_{\mathrm{I}}$ | No. of fully contagious Erlang states | 16 |
| $n_{\mathrm{L}}$ | No. of late-infectious Erlang states | 16 |
| $D_E$ | Average duration of latency period | 3.7 days |
| $D_P$ | Average duration of prodromal period | 1 day |
| $D_I$ | Average duration of fully contagious period | 7 days |
| $D_L$ | Average duration of late infectious period | 7 days |
| $\varepsilon$ | Transition rate of latent states | $n_{\mathrm{E}}/D_E$ |
| $\varphi$ | Transition rate of prodromal states | $n_{\mathrm{P}}/D_P$ |
| $\gamma$ | Transition rate of early infectious states | $n_{\mathrm{I}}/D_I$ |
| $\delta$ | Transition rate of late-infectious states | $n_{\mathrm{L}}/D_L$ |
| $\alpha$ | Transition rate from transient multi infections to multi-infected states | 3.2/day |
| $f_{\mathrm{Sick}}$ | Fraction of symptomatic (sick) infections | 58% |
| $\tilde{f}_{\mathrm{Sick}}$ | Fraction of sympt. multi infections in fully contagious & late inf. periods | 64.4% |
| $f_{\mathrm{Iso}}$ | Fraction of single-infected (sick) who are isolated (or home isolated) | 0.5 |
| $\tilde{f}_{\mathrm{Iso}}$ | Fraction of multi-infected (sick) who are isolated (or home isolated) | 0.55 |
| $f_{\mathrm{Dead}}$ | Fraction of single-infected (sick) who die from the disease | 3% |
| $\tilde{f}_{\mathrm{Dead}}$ | Fraction of multi-infected (sick) who die from the disease | 4% |

Summary of parameters describing number of Erlang states, durations, transition rates, morbidity and mortality, and their default parameter choices.
