## Supplementary material for "Is increased mortality by multiple exposures to COVID-19 an overseen factor when aiming for herd immunity?": Table S3

**S3 Table.** Summary of model parameters.

| Parameter | Definition | Value/Eq. |
|---|---|---|
| $\lambda(t)$ | Total force of infection | $\lambda^{(\mathrm{s})}(t) + \lambda^{(\mathrm{m})}(t)$ |
| $\lambda^{(\mathrm{s})}(t)$ | Force of infection leading to single infection | Eq. (22) |
| $\lambda^{(\mathrm{m})}(t)$ | Force of infection leading to multi infections | Eq. (23) |
| $\lambda^{(\mathrm{s})}_{\mathrm{Ext}}$ | External force of infection leading to single infections | 50/day |
| $\lambda^{(\mathrm{m})}_{\mathrm{Ext}}$ | External force of infection leading to multi infections | 0 |
| $\tilde{m}$ | Fraction of contacts with multi infected that cause multi infections | 0.2 |
| $q_E$ | Prob. that single-infected lead to transient multi infection in latent states | 1 |
| $q_P$ | Prob. that single-infected lead to transient multi infection in prodromal states | 0.75 |
| $q_I$ | Prob. that single-infected lead to transient multi infection in fully contagious states | 0.5 |
| $q_L$ | Prob. that single-infected lead to transient multi infection in late-infectious states | 0.25 |
| $\bar{R}_0$ | Annual average basic reproduction number | 2.5 |
| $a$ | Amplitude of the seasonal fluctuation of the basic reproduction number | 0.43 |
| $t_{R_{0_{\max}}}$ | Day when $R_0$ reaches its maximum | 270 |
| $c_P$ | Relative contagiousness in the prodromal period | 0.5 |
| $c_I$ | Relative contagiousness in the fully contagious period | 1 |
| $c_L$ | Relative contagiousness in the late infectious period | 0.25 |
| $\beta_P(t)$ | Seasonally varying effective contact rate of prodromal ind. | Eq. (18) |
| $\beta_I(t)$ | Seasonally varying effective contact rate of full contagious ind. | Eq. (19) |
| $\beta_L(t)$ | Seasonally varying effective contact rate of late-infectious ind. | Eq. (20) |

Summary of parameters describing infectiousness, contact rates, forces of infection, and their default parameter choices.
