## Supplementary material for "Is increased mortality by multiple exposures to COVID-19 an overseen factor when aiming for herd immunity?": Table S4

**S4 Table.** Summary of model parameters.

| Parameter | Definition | Value |
|---|---|---|
| $Q_{\mathrm{max}}$ | Maximum capacity of the isolation units | 1/100 |
| $p_{\mathrm{Home}}$ | Prevented fraction of contacts of individuals who are isolated at home | 0.75 |
| $p_{\mathrm{Dist}}$ | Prevented fraction of contacts because of general social-distancing measures | 0.4 |
| $t_{\mathrm{Iso}_1}$ | Day when the case-isolation measures start in the population | day 0 |
| $t_{\mathrm{Iso}_2}$ | Day when the case-isolation measures end in the population | day 365 |
| $t_{\mathrm{Dist}_1}$ | Day when social-distancing measures start | day 0 |
| $t_{\mathrm{Dist}_2}$ | Day when social-distancing measures end | day 45 |

Summary of parameters describing interventions and default parameter choices.
