## Supplementary material for "Is increased mortality by multiple exposures to COVID-19 an overseen factor when aiming for herd immunity?": Fig S1

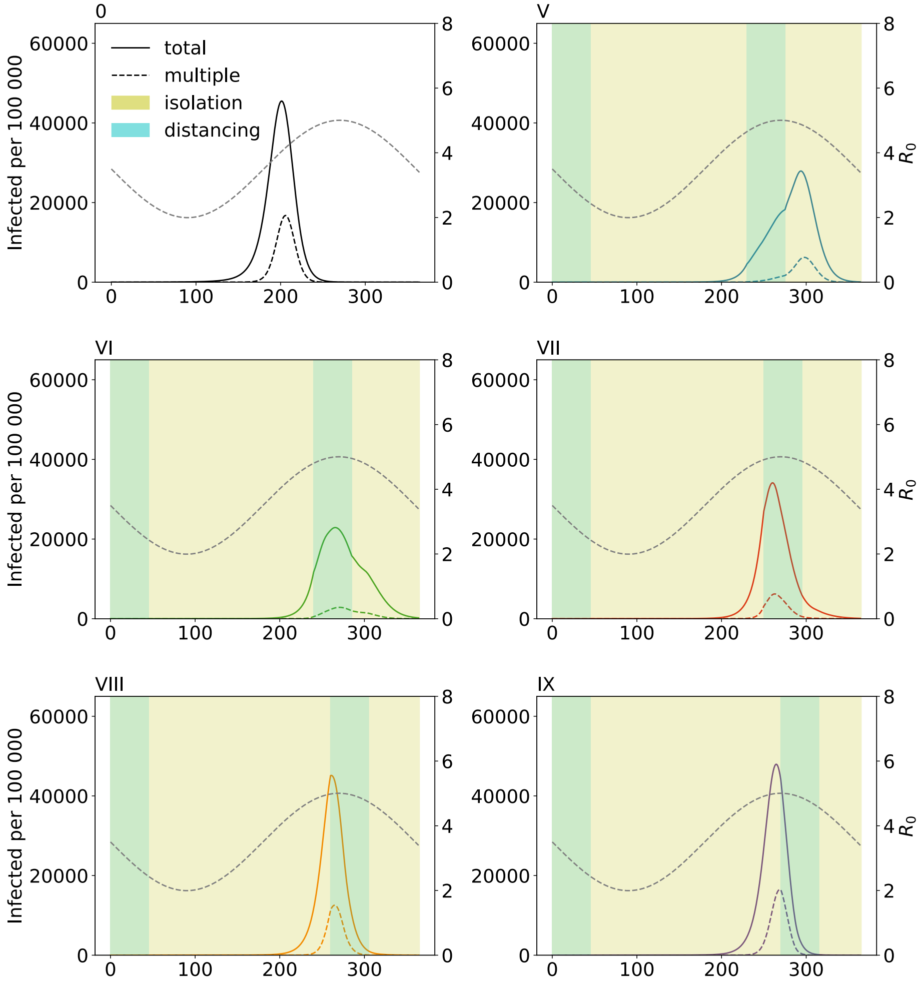


**S1 Fig. Implications of a second lockdown.** Shown are the numbers of infected (including latent, prodromal, fully contagious, and late infectious) single and multi infected individuals (solid) and multi infected individuals only (dashed lines). Seasonally vary $R_0$ is indicated by the gray dashed line corresponding to the y-axis on the right-hand side. Different panels correspond to scenarios described in Table 2. Periods during which case isolation is sustained are indicated in yellow and periods of lockdown are indicated in cyan (overlapping the periods of case isolation). Parameters are given in S1 Table – S4 Table.
