## Supplementary material for "Is increased mortality by multiple exposures to COVID-19 an overseen factor when aiming for herd immunity?": Fig S2

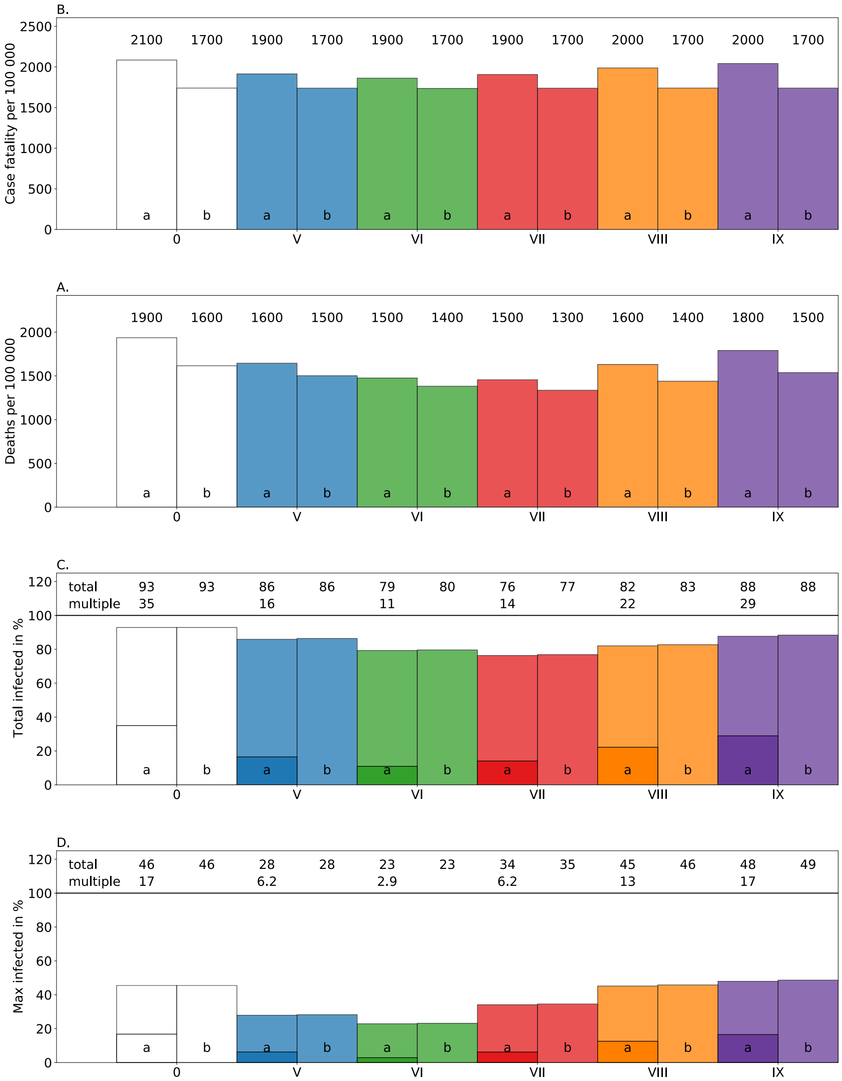


**S2 Fig. Implication of a second lockdown.** For each scenario (0, V – IX), values are given for the assumption that multi infections cause higher morbidity and mortality and are more likely to be isolated than single infections as described in the text (a) and for the assumption that morbidity, mortality and isolation are the same as for single infection (b). Shown are the total number of deaths (A); (B) case fatality (i.e., deaths per infected individuals); (C) total percentage (of population) being infected or multi infected; and (D) the maximal number of individuals infected or multi infected at the epidemic peak, for scenarios as specified in Table 2 contrasting the case with increased morbidity and mortality of multi infections (a) and no effect of multi infections on mortality and morbidity (b), i.e., the case in which single and multi infections are indistinguishable. The horizontal lines in scenario (a) in (C) and (D) indicate the number of multi infections characterized by increased morbidity and mortality given by (33) in S1 Appendix. Parameters are given in S1 Table – S4 Table.
